# Superspreading k-cores at the center of COVID-19 pandemic persistence

**DOI:** 10.1101/2020.08.12.20173476

**Authors:** Matteo Serafino, Higor S. Monteiro, Shaojun Luo, Saulo D. S. Reis, Carles Igual, Antonio S. Lima Neto, Matías Travizano, José S. Andrade, Hernán A. Makse

## Abstract

The spread of COVID-19 caused by the recently discovered SARS-CoV-2 virus has become a worldwide problem with devastating consequences. To slow down the spread of the pandemic, mass quarantines have been implemented globally, provoking further social and economic disruptions. Here, we address this problem by implementing a large-scale contact tracing network analysis to find the optimal quarantine protocol to dismantle the chain of transmission of coronavirus with minimal disruptions to society. We track billions of anonymized GPS human mobility datapoints from a compilation of hundreds of mobile apps deployed in Latin America to monitor the evolution of the contact network of disease transmission before and after the confinements. As a consequence of the lockdowns, people’s mobility across the region decreases by ~53%, which results in a drastic disintegration of the transmission network by ~90%. However, this disintegration did not halt the spreading of the disease. Our analysis indicates that superspreading k-core structures persist in the transmission network to prolong the pandemic. Once the k-cores are identified, the optimal strategy to break the chain of transmission is to quarantine a minimal number of ’weak links’ with high betweenness centrality connecting the large k-cores. Our results demonstrate the effectiveness of an optimal tracing strategy to halt the pandemic. As countries race to build and deploy contact tracing apps, our results could turn into a valuable resource to help deploy protocols with minimized disruptions.

---

In the absence of vaccine or treatment for COVID-19, state-sponsored lockdowns have been implemented worldwide to halt the spread of the ongoing pandemic creating large social and economic disruptions [1–3]. In addition, some countries have also implemented digital contact tracing protocols to track the contacts of infected people and reinforce quarantines by targeting those at high risk of becoming infected [4–7]. Here we develop, calibrate, and deploy a contact tracing algorithm to track the chain of disease transmission across society. We then search for intelligent quarantine protocols to halt the epidemic spreading with minimal social disruptions [8–13].

Our study uses two complementary datasets. The first includes data from ’Grandata-United Nations Development Programme partnership to combat COVID-19 with data’ [14]. It is composed of anonymized global positioning system (GPS) data from a compilation of hundreds of mobile applications (apps) across Latin America that allow to track the trajectories of people (users). The data identify each mobile phone device with a unique encrypted mobile ID and specifies its latitude and longitude location through time, encoded by geohash with 12 digits precision. Typically, this dataset generates ~ 450 million data points of GPS location per day across Latin America in particular in teh state of Ceará, Brazil (see Methods).

The second dataset is an anonymized list of confirmed COVID-19 patients obtained from the Health Department authorities from both states. It includes the geohash of the address, the SARS-COV-2 test detection date and first day of symptoms of COVID-19. We cross-match the geolocation of the patients with the GPS dataset obtaining the encrypted mobile ID of the patients (see Methods). We then trace the geolocalized trajectories of COVID-19 patients during a period -14/+7 days from the onset of symptoms to look for contacts of the infected person to define the transmission network using the model described below.

## COVID-19 model

The COVID-19 spreading model is represented by a Susceptible-Exposed-Infectious-Recovered (SEIR) process [9] (Fig. 1a). Infectiousness starts 2 days before and lasts up to 5 days after the onset of symptoms [15]. In this paper, we add two days to each of these limits to conservatively capture most transmissions. Thus, in principle, to trace those people potentially infected by COVID-19 patients, we track contacts 4 days before and 7 days after the reported date of first symptoms (see Fig. 1a). In addition, we extend the tracing period back in time to also consider exposures that could come from asymptomatic cases. Exposures start the incubation period of the infected person which can occur up to 12.5 days before onset of symptoms (5.2 days on average, 95% percentile 12.5 days [16, 17], Fig. 1a). To conservatively trace exposure events, we add ~2 days to this incubation period and obtain the widely used 14 days period. Hence, to trace transmission and exposure cases, we perform contact tracing over -14/+7 days from onset of symptoms (Fig. 1a). We note that the peak of infectiousness as well as 44% (95% confidence interval, 25-69%) of infected cases occur during the pre-symptomatic stage [15]. Thus, performing contact tracing is essential to stop the spreading disease.

**FIG. 1.**
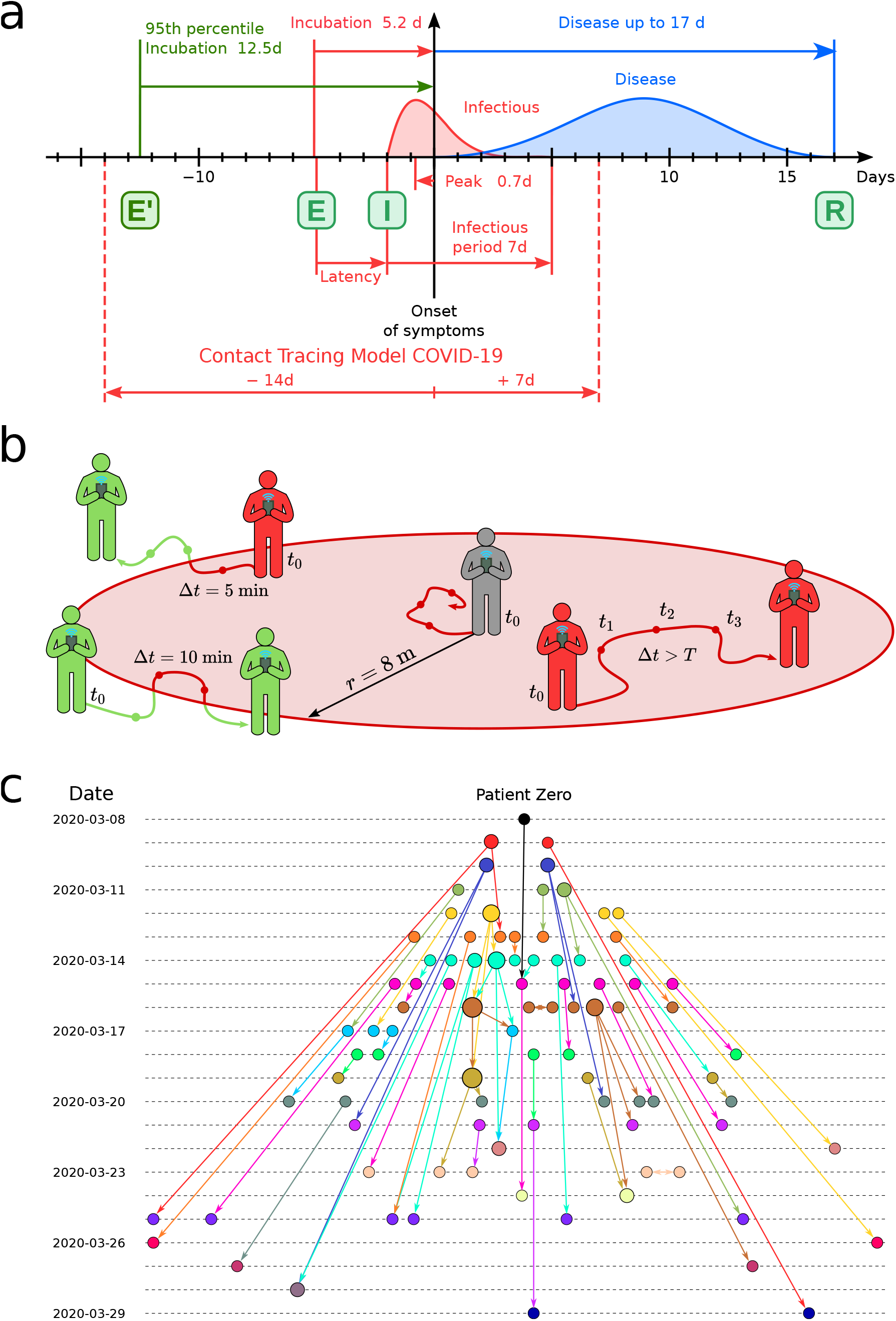
COVID-19 contact model. **(a)** Infectiousness profile of COVID-19. The COVID-19 pandemic is represented with a SEIR model. From exposure (E) the virus is incubated in average for 5.2 days (12.5 days 95^th^ percentile), starting the symptoms 2 days after infectiousness (I) and lasting the disease up to 17 days to recover (R). We use a window -14/+7 days from the first symptoms to detect infectious and exposure. **(b)** Contact area used in the contact tracing model. The grey person is at the first datapoint of the source at *t*_0_. We collect all datapoints for every user in a *T*=30 min forward window (*t*_1_, *t*_2_, *t*_3_, …, *t*_0_+*T*) within an 8 m circle from the initial position. For each target (green and red) we compute the average position and the time spent inside the contact area (red part of the trajectory line). **(c)** Partial transmission tree of outbreak of confirmed SARS-CoV-2 infection identified by contact tracing during calibration in the month of March 2020. Links goes from the source of infection to the target. The colors represent the day of first symptoms for each node and size is the out-degree.

## Contact model

The GPS geolocation of the trajectories of both infected and susceptible people is used to trace several layers of contacts in the transmission network using the following model. A contact at time stamp *n* is initiated with an infected user (source) at time *t*_0_ (see Fig. 1b). At *t*_0_ we draw a contact area as a circle centered in the source position with a radius *r*. We then gather all the GPS datapoints from susceptible users (targets) that enter the contact area from *t*_0_ to *t*_0_ + *T*, where *T* is the total exposure time. We follow the trajectories of source and target within the time-space area and compute the probability of infection at time stamp n as *p_i_*[*n*] = *p_d_*[*n*] · *p_t_*[*n*], where *p_d_*[*n*] is the spatial component, and *p_t_*[*n*] is the temporal component. When the average overlap between source and target is zero, then *p_d_*[*n*] = 1, and when the overlap is 2*r*, then *p_d_*[*n*] = 0. On the other hand, when the exposure time ≥ *T*, then *p_t_*[*n*] = 1, and decreases to *p_t_*[*n*] = 0 as the exposure time decreases (see Methods for definitions). The probability *p_d_*[*n*] quantifies the contact probability for two users in the same area defined by *r*. A contact requires non only a space overlapping but also a time overlap, *p_t_*[*n*], which quantifies the probability that two users met based on the time commonly spent in the same area. We then combine these two probabilities for each timestamp *n* into their product.

Contacts with low probability of infection *p_i_*[*n*], but repeated throughout time, can also infect the target. To incorporate this effect in the model, we define the probability of infection for a series of repeated contacts *P_i_*[*n*] as a recursive formula from time 1 to *n* with *P_i_*[0] = 0:

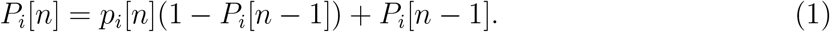

The iteration of contacts between source and target, *P_i_*[*n*], generates higher probability of infection than a single contact *p_i_*[*n*]. This means that there is a difference between a short single contact between two people and short repeated contacts between the same people. The latter scenario should have a larger probability than the former to become infected.

While the distribution of *p_i_*[*n*] is homogenous without a clear threshold for an infectious contact, *P_i_*[*n*] presents a very polarized distribution where the values are accumulated in the extremes: *P_i_* = 0 or *P_i_* = 1 (see SI Fig. 11a). Thus, *P_i_*[*n*] is better indicator than *p_i_*[*n*] to separate infectious from non-infectious contacts. A contact is then considered infectious when this probability exceeds a certain threshold, *P_i_*[*n*] > *p_c_*. The hyperparameters of the contact model (*T*, *r*, *p_c_*) are obtained by calibrating the model using only the contacts between infected people to reproduce the basic reproduction number *R*_0_ = 2.78 in Ceará in the month of March, 2020 (see Methods). We obtain *T* = 30 min, *r* = 8 m and *p_c_* = 0.9. Thus, a contact is defined with probability one when exposure is at least 30 minutes within a distance ≪ 8m. This calibration procedure provides the partial transmission tree of the outbreak from patient zero to the end of the calibration period shown in Fig. 1c.

## Transmission network model

Next, we create the contact network of coronavirus transmission by first tracing the trajectories of confirmed COVID-19 patients to search for contacts -14/+7 days from the onset of symptoms using the above model. From the first contact layer, we add four layers of contacts to constitute the contact network of transmission that is used to monitor the progression of the pandemic. The time-varying network is aggregated to a snapshot defined over a time window of a week [9]. We find that other aggregation windows give similar results as presented.

Next, we analyze the spatio-temporal properties of the contact network. The government of the State of Ceará imposed a mass quarantine on March 19, 2020 which led to a decrease in people’s mobility by 56.5% as shown in Fig. 2a. During the lockdown, only the displacements of essential workers were allowed. A large decrease in mobility is also observed across all Latin America, see [14].

**FIG. 2.**
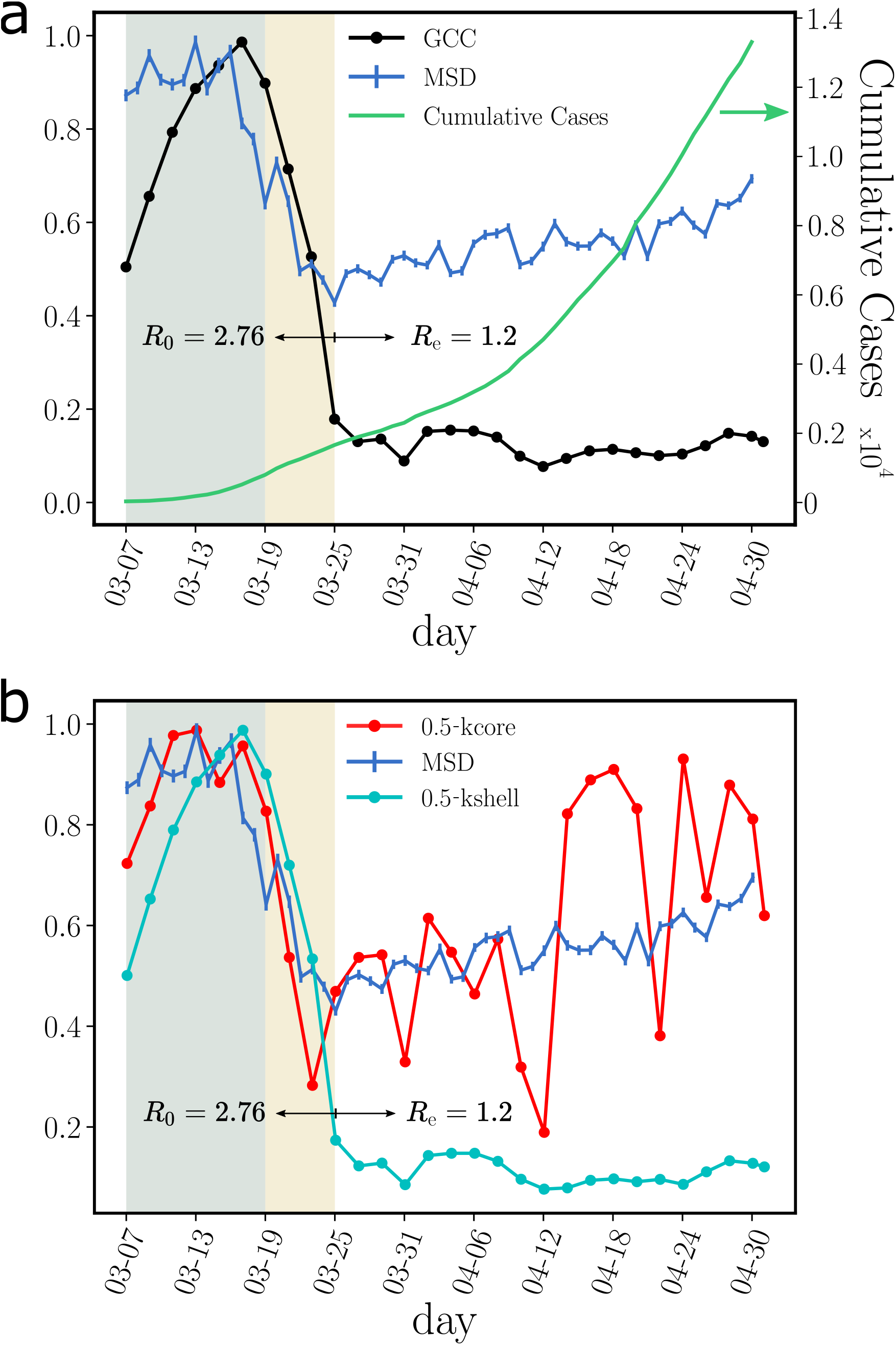
Structural components of transmission networks across the lockdown. **(a)** Evolution for different metrics in Ceará, Brazil, previous to the mass quarantine (grey area), right after the imposed quarantine (yellow area) and later. The plot shows the root mean square displacement (MSD) normalized by the maximum value over the total period (blue), the cumulative number of cases (green) and the size of the GCC normalized by the maximum value over the total period (black). The uncertainty corresponds to the standard error (SE). The mobility data is showcased in the Grandata-United Nations Development Programme map shown in https://covid.grandata.com. The initial rise in GCC is due to the lack of data before March 1. **(b)** The plot shows the 0.5-kcore size (red), the 0.5-kshell size (cyan) all normalized by their respective maximum value pre-lockdown. While the size of the 0.5-kshell is reduced drastically during the lockdown, the 0.5-kcore was not reduced as much and keeps increasing, contributing to sustain the pandemic. The 0.5-kcore seems to follow the trend in the MSD, which we plot again to show this trend.

## Giant connected component (GCC)

To understand the effect of the lockdown on the contact network, we think by analogy with a ’bond percolation’ process [9, 10, 18]. In bond percolation, the network connectivity is reduced by removing a small fraction of links (bonds) between nodes, and the global disruption in network connectivity is monitored by studying the normalized size of the giant connected component (see Methods). Following this analogy, the lockdown acts as a percolation process, and therefore we monitor the GCC of the transmission network before and after the lockdown. We find a drastic percolation transition [9, 18] within 6 days of the implementation of the lockdown on March 19, when the GCC is almost fully dismantled decreasing by 89.6% of its pre-lockdown size (Fig. 2a).

Despite the disintegration of the GCC, the cumulative number of cases kept growing albeit at a lower rate (Fig. 2a). We find that the mass quarantine was able to reduce the basic reproduction number from *R*_0_ = 2.78 before lockdown to an effective reproduction number of *R_e_* = 1.2 after the lockdown (Fig. 2a). Despite this disruption in the network connectivity, *R_e_* has not decreased below one, as it would have been needed to curb the spread of the disease.

The drastic reduction in the GCC is visually apparent in the contact networks in Fig. 3. Before lockdown on March 19 (Fig. 3a), the network is a strongly-connected unstructured ’hairball’. Eight days into the lockdown on March 27 (Fig. 3b), the network has been untangled into a set of strongly-connected modules integrated by tenuous paths of contacts. This structure is even more pronounced a few weeks later on April 28 (Fig. 3c).

**FIG. 3.**
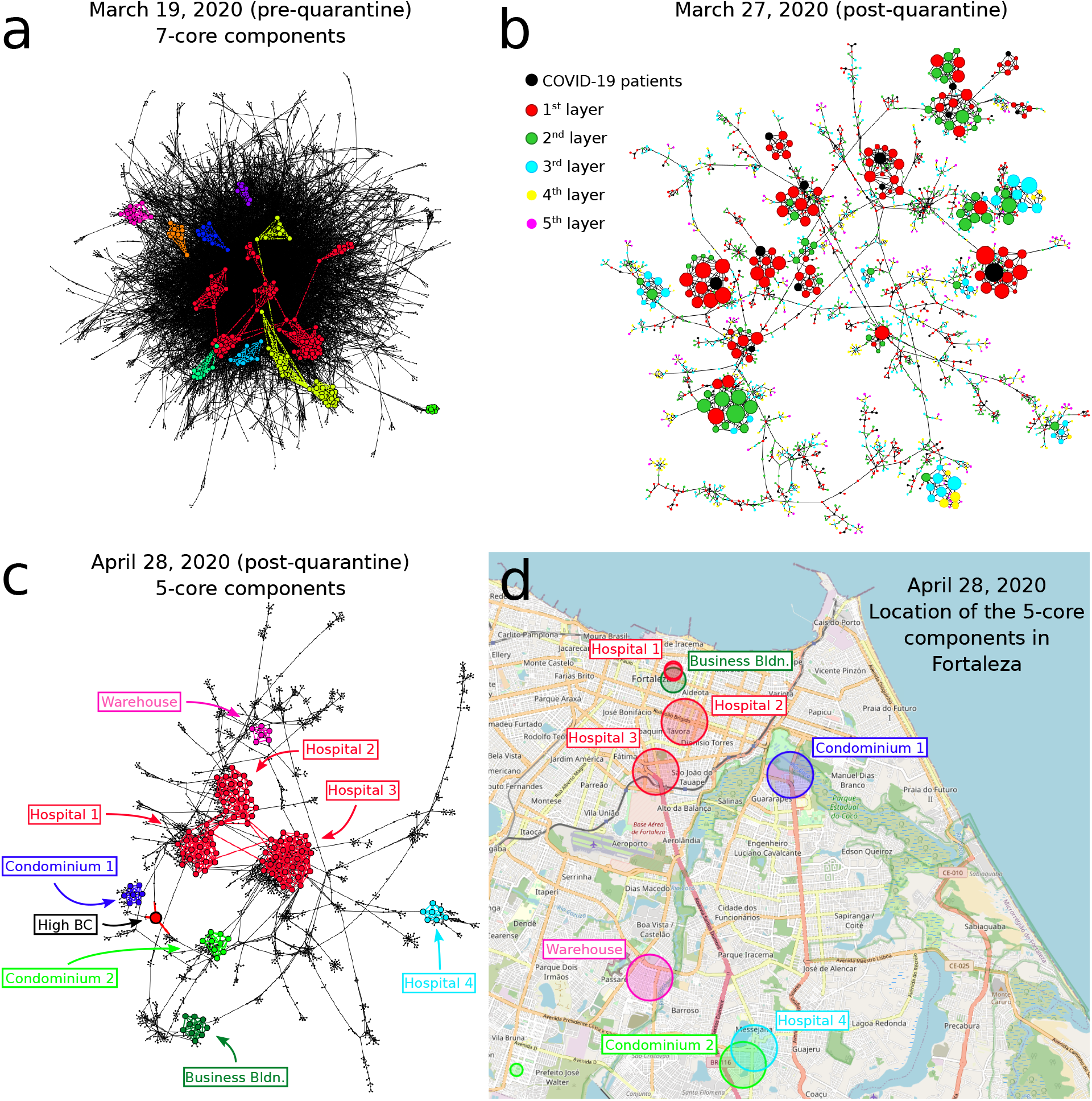
Evolution of GCC and k-cores over the quarantine. Disease transmission networks in the state of Ceará over time before and after the lockdown on March 19, 2020. **(a)** Transmission network on March 19 (pre-lockdown). A hairball highly-connected network is observed. The disconnected components of the 7-core (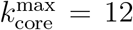 in this network) are colored. These components are well connected into the hairball network as expected since mobility and connectivity is high. **(b)** The pre-quarantine hairball in **(a)** has been untangled and the k-cores have emerged 8 days into the lockdown on March 27. Here, we color the nodes according to layers of the transmission network starting at COVID-19 patient (black nodes). Size of nodes is according degree. **(c)** Network on April 28 including the components of the 5-core in different colors (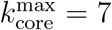 for this network). Visible is the high betweenness centrality node representing the weak-link of this k-core. **(d)** We plot the location of the contacts in the map of Fortaleza constituting the components of the 5-core of the April 28 in **(c)**. The size of the circles in the map corresponds to the number of contacts inside each location. The colors correspond to the clusters of the 5-core in **(c)**. The 5-core sustaining transmission is composed of clusters of contacts localized in hospitals, large warehouses and business buildings. Hospital 3, one of the largest in Fortaleza, constitutes the maximal (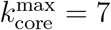 of the pandemic.

## Superspreading k-core structures

The highly connected modules found in Fig. 3b and 3c are k-core structures [19–22] of higher complexity than the GCC (which is a 1-core), that are known to sustain an outbreak even when the GCC has been disintegrated [9, 22]. The k-core of a graph is the maximal subgraph in which all nodes have a degree (number of connections) larger or equal than *k* [19–22]. The k-shell is the periphery of the k-core and is composed by all the nodes that belong to the k-core but not to the (k+1)-core (see Methods for definitions and SI Figs. 12, 13, and 14). The k-core is obtained by iteratively pruning the nodes with degree smaller than *k*. For instance, the 3-core is obtained by removing the 1-shell and 2-shell in a k-shell decomposition process (see SI Figs. 12, 13). Thus, all nodes in a k-core have at least degree *k*, and are connected to other nodes with degree at least *k* too. K-cores are nested and can be made of disconnected components (see SI Fig. 14). High k-cores are those with large *k* up to a maximal 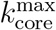, and constitute the inner most important part of the network. In theory, the high k-cores are known from network science studies to be the reservoir of disease transmission persistence [9, 22]. On the contrary, low peripheral k-shells (see SI Fig. 12) do not contribute as much to the spread as the high inner k-cores.

Figure 2b shows that despite the disappearance of the GCC, there is a significant maximal k-core that was not dismantled by the mass quarantine. The figure shows that the outer k-shells of the transmission network (i.e., the 0.5-kshell defined as the union of the k-shells with 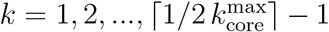, see Methods) are disintegrated in the lockdown, decreasing by 91% with respect to their pre-quarantine size, in tandem with the GCC. However, the inner k-core (i.e., the 0.5-kcore defined as the k-core with 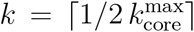, see Methods) persists in the lockdown. The figure shows that the decrease of the 0.5-kcore is only 50% compared to the 91% decrease of the 0.5-kshell; the former even increases slightly at the end of April, following the same trend in mobility (see Fig. 2b). This process is visually corroborated in the evolution of the networks seen from Fig. 3a to 3c where we observe the disappearance of the peripheral k-shells and the persistence of the maximal k-core. Indeed, the unessential contacts in the peripheral k-shells may have been first pruned during social distancing.

Using numerical simulations, we corroborate that the infection can persist in these high k-cores of the network while virus persistence in outer k-shells is less important [9, 22]. We use a SIR model on the transmission network (Fig. 4a and SI Fig. 15a) showing that the maximal k-cores of the network sustain the spreading of the disease more efficiently than the outer k-shells. Thus, the maximal k-core components of the contact network are plausible drivers of disease transmission. Apart from this structural explanation (i.e., k-core), epidemiological factors may also play a role in the persistence of the disease, such as a transition of the disease to vulnerable communities with high demographic density, or with large inhabitants per household where isolation is poorly fulfilled.

**FIG. 4.**
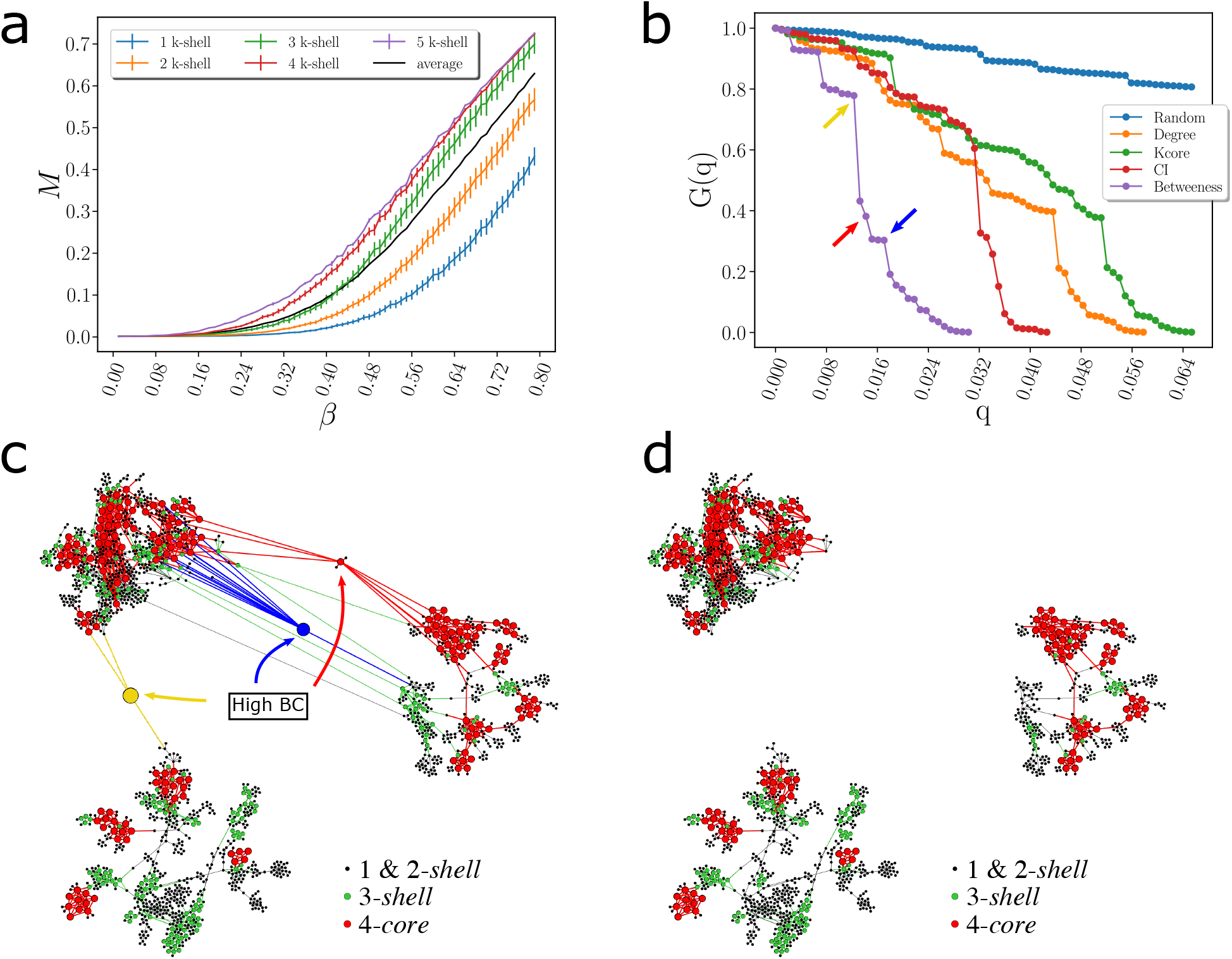
Weak links and k-cores. **(a)** Average size of infected population, M [22], in an outbreak average over all starting nodes in a k-shell as a function of the probability of infection *β* for a SIR model on the network in Fig. 3c during the lockdown. The black is the average value over all the network. The average divides the k-shell contribution to the spreading of the virus in two groups: above and below the average. The 0.5-kcores have maximal spreading and the 0.5-kshell have minimal spreading. Error bars correspond to SE. **(b)** Optimal percolation analysis performed over the network in Fig. 3c during the lockdown in following different attack strategies and their effect on the size of the largest connected component *G*(*q*) versus the removal node fraction, *q*. Nodes are removed (in order of increasing efficiency): randomly (blue); by the highest k-shell followed by high degree inside the k-shell [22]; by highest degree (orange); by collective influence (red) [13]; and by the highest value of betweenness centrality (green) [26, 27]. After each removal we re-compute all metrics. The most optimal strategy is removing the nodes by the highest value of betweenness centrality. **(c)**-**(d)** Effect of removing three high betweenness centrality nodes shown in Fig. 4b in the network of Fig. 3c. **(c)** We show the 2-core component of the network after the removal of 12 high betweenness centrality nodes. The red node is the one with the highest betweenness centrality value (next node to remove, 13th) and the blue node is the 14th removal. Different k-cores and k-shell are in different colors. **(d)** Network k-cores are disintegrated after the removal of the high BD nodes.

When we plot the geolocation of the contacts forming the maximal k-core in the map of Ceará, we find that these contacts take place in highly transited areas of the capital Fortaleza, such as hospitals, business buildings, warehouses as well as large condominiums, see Fig. 3d. These contacts generate superspreading k-core events that generalize the conventional notion of superspreaders, which refer mainly to individuals with large number of transmission contacts [23–25]. However, connections are not everything [11, 12]. K-core superspreaders not only generate a large number of transmission contacts, but their contacts are also highly connected people, and so forth.

## Optimal quarantine

The existence of k-cores in the transmission network suggests that a more structured quarantine could be deployed to either isolate or destroy those cores that help maintain the spread of the virus. We perform an optimal percolation analysis [11–13] to find the minimal number of people necessary to quarantine that will dismantle the transmission network. We follow different strategies to find the optimal breakdown of the network by ranking the nodes based on (1) the number of contacts (hub-removal) [9, 11, 12], (2) the largest k-shells and then by the degree inside the k-shells [9, 22], (3) the collective influence algorithm for optimal percolation [13], and (4) betweenness centrality [26–29] (we also try other centralities, see Methods for definitions).

Figure 4b shows the normalized size of the GCC versus the fraction of removal nodes following different strategies, as well as a random null model of removal in a typical network under lockdown in April 28 (March 19 pre-lockdown results are plotted in SI Fig. 15b). While the disease can persist in the k-cores (Fig. 4a), quarantining people directly inside the maximal k-core is not an optimal strategy. The reason is that k-cores are populated by hyper-connected hubs that requiere many removals to break the GCC [28] (around 7%, see Fig. 4b). For the same reason, removing directly the hubs is not the optimal strategy either, since the hubs are within the maximal k-core and not outside. A collective influence strategy [13] improves over hub-removal since it takes into account how hubs are spatially distributed, yet, it is far from optimal. Clearly, Fig. 4b shows that the best strategy is to quarantine people by their betweenness centrality. By removing just the top 1.6-2% of the high betweenness centrality people, the GCC is disintegrated. This is consistent with the particular structure of the transmission networks seen in Fig. 3b, c and Fig. 4.

The betweenness centrality of a node is proportional to the number of shortest paths in the network going through that node. Thus, given the particular structure of the networks in Figs. 3b, c, and Fig. 4c, the high betweenness centrality nodes are the bottlenecks of the network, i.e., loosely-connected bridges between the largely-connected k-cores components. These connectors are the celebrated ’weak links’, fundamental concept in sociology proposed by Granovetter [30], according to which, strong ties (i.e., contacts in the k-cores) clump together forming clusters. A strategically located weak tie between these densely ’knit clumps’, then becomes the crucial bridge that transmits the disease (or information [30]) between k-cores. These weak links are people traveling among the different k-cores components allowing the disease to escape the cores into the rest of society. These bridges are displayed in the network of Fig. 4c as yellow, blue and red nodes. The removal of these high betweenness centrality people disconnects the k-core components of the network entirely, as shown in Fig. 4d, halting the disease transmission from one core to the other [28, 31].

An important finding is that quarantining the large superspreading k-cores is neither optimal (as shown in Fig. 4b, green curve) nor practical, since they are mainly comprised by chiefly essential workers who need to remain operational (Fig. 3d). Thus, the best strategy, in conjunction with the mass quarantine, is then to disconnect these k-cores from the rest of the social network (Figs. 4c and 4d), rather than quarantining the people inside the k-cores. This can be performed by quarantining the high betweenness centrality weak-links that simultaneously preserve the operational k-cores. However, individuals belonging to the maximal k-cores should be tested at a higher frequency to promptly detect their infectiousness before the symptoms start, to help control the spreading inside the k-cores.

## Summary

Isolating the k-core structures by quarantining the high betweenness centrality weak links proves to be the most effective way to dismantle the GCC of the disease while keeping essential k-cores working. While destroying the strong links and cores is a less manageable task to execute and control, isolating the weak links between cores is a more feasible task that will assure the dismantling of the GCC. In other words, if one core is infected, the disease will be controlled within that core and not extended to the rest of society.

As governments around the world are racing to roll out digital contact tracing apps to curb the spread of coronavirus [4, 5], our modeling suggests possible intelligent quarantine protocols that could become key in the second phase of reopening economies across the world and, in particular, in developing countries where resources are scarce. SI Section I addresses issues of sampling bias and coverage of the data relevant for implementation. Overall, our network-based optimized protocol is reproducible in any setting and could become an efficient solution to halt the critical progress of the COVID-19 pandemic worldwide drawing upon effective quarantines with minimal disruptions.

## Data Availability

We make available the anonymized geolocation GPS data used in the present study in the States of Ceará under request to M. Travizano at. Source codes of all algorithms
used in this study are available at https://github.com/makselab/COVID19 and https://github.com/makselab/COVID19_contact_detector.

## Acknowledgments

We are grateful to S. Alarcón-Díaz, N. Della Penna, and I. Be-lausteguigoitia. We also thank J. Maciel, G. Sousa, J. A. P. Barreto, and R. Sousa from the Health Secretariat of Fortaleza for data curation.

## Authors contributions

HAM designed the project. All authors contributed to analysis and writing of the paper.

## Data and code availability

This study was approved by the Institutional Review Board (IRB) at City College of New York and Universidade Federal do Ceará. Patient data was used with the approval and consent obtained by the Epidemiological Surveillance Department, Fortaleza Health Secretariat at the Prefeitura de Fortaleza, Ceará, Brazil. In the context of the ongoing health crisis, we make available the anonymized geolocation GPS data used in the present study in the States of Ceará under request to M. Travizano at. Source codes of all algorithms used in this study are available at https://github.com/makselab/COVID19 and https://github.com/makselab/COVID19_contact_detector.

## METHODS

### Datasets

The GPS dataset from ’Grandata-United Nations Development Programme partnership to combat COVID-19 with data’ https://covid.grandata.com [14] provides the geolocation data of users of a compilation of hundreds of mobile apps across Latin America. Each datapoint registers the geohash location of a mobile ID with a 12 digits precision (cm resolution, although we use meter resolution due to noise in the determination of the exact GPS location). The mobile ID is MD5 hashed data associated with a hash of the MADID (Mobile Advertising ID).

**TABLE I.** GPS datasets of Ceará and Puebla from [14].

|  | Ceará |  |  |
| --- | --- | --- | --- |
| Month 2020 | Unique users | Daily users | GPS Datapoints |
| March | 111,168 | 43,231 | 153,282,989 |
| April | 145,148 | 49,913 | 161,304,623 |
| May | 158,405 | 63,489 | 221,381,875 |
| Total | 257,606 | 52,211 | 535,969,487 |

See [14]: The full GPS dataset covers 10 countries across Latin America: Argentina, Brazil, Chile, Colombia, Ecuador, Guatemala, Mexico, Paraguay, Peru and Uruguay. Average number of users in the dataset per day: Uruguay: tens of thousands of unique users per day. Argentina: hundreds of thousands of unique users per day. Mexico: Millions of unique users per day. Brazil: tens of millions of unique users per day. Over the month of March, lockdowns have been imposed on each country. Pre-quarantine dates in Latin America range: March: 1st through 19th, 2020. Quarantine in Latin America dates range: March 20th till today, as of June 9.

We measure the average root mean square displacement (MSD) of the users in each country per day. Averaging over all the countries in Latin America we measure a -53.4% reduction in the MSD from a reference date before lockdown on March 6, 2020 to May 1, 2020, see [14].

In the state of Ceará, we find a reduction in mobility from MSD=988m pre-quarantine to MSD= 430m, giving a reduction of 56.5%. The GCC of transmission contacts defined over one week window in the state of Ceará is reduced from a size of 9402 users pre-lockdown to an average size of 983 users per week during the lockdown, giving a reduction of the GCC to -10.4% from its original pre-quarantine size.

Patient’s datasets are provided by the Department of Epidemiological Surveillance of the Fortaleza Health Secretariat in the city of Fortaleza, Ceará, Brazil. The research was approved by IRB at CCNY and UFC. Patient data was used with the approval and consent from the Epidemiological Surveillance Department, Fortaleza Health Secretariat at the Prefeitura de Fortaleza, Ceará, Brazil.

We cross the information from the Health Department dataset with the GPS dataset detecting the mobile ID’s that are related to any confirmed case from the Health Department data using their geolocalized address. We set the night time period from 10 PM to 5 AM as a time window with high probability of being at the location. Afterwards, we identify the mobile ID of the user that spent more than 15% of the total time in the location in those time-spatial areas during March-May 2020. We assign that mobile ID to the patient.

In the Fortaleza dataset there were 8,323 cumulative cases from March 1 to May 7, 2020, out of which 5,814 contain geolocation and date of first symptoms, and 1,440 cases were matched to the GPS dataset. Lockdown started on March 19, 2020. On the month of March, 465 cases are matched which are used to calibrate the model. The number of nodes in the transmission tree of Fig. 1c is 90, the number of connections in the tree is 60, and the number of secondary infections in the tree is 52. Our results can be applied to other areas as well. In preliminary studies we find that they are reproducible in other studied regions such as the state of Puebla, México.

### Digital contact tracing model

For each time stamp starting at *t*_0_ we draw the time-spatial area shown in Fig. 1b and collect all the geolocalized data points inside the area. Afterwards we compute the average position for each user and the interval of time they have been within the contact area: 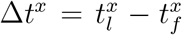, where 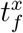 and 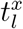 are the first and last data points a user *x* (target (*t*) or source (*s*)) was within the circle.

The first component of the probability of infection is the space component [10]:

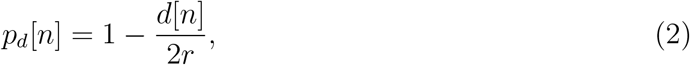

with *d*[*n*] = |〈*s*〉 − 〈*t*〉|, where 〈*x*〉 in {*s*, *t*} refers to the average position of *x* inside the time-space area. That is, it is the distance between the source and target average positions of the data points within the contact area.

The time component is proportional to the overlapped amount of time that source and target spent within the contact area:

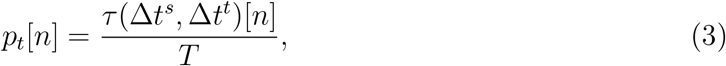

and

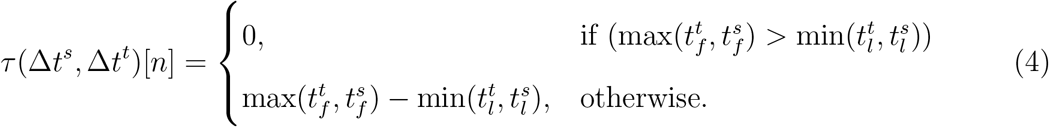

Here, *τ*(Δ*t^s^*, Δ*t_t_*)[*n*] is the overlapped time in the time stamp *n*. Note that each user needs two data points within the contact area to define its Δ*t^x^* and *τ*[*n*], otherwise *τ*[*n*] = 0. When the source is the one that does not fulfill this requirement we omit that contact area.

Using both components, we define the single contact probability:

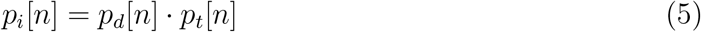

for each time stamp *n*. Using the recursive Eq. (1) we compute a unique value for each contact accumulating recursively the values of all single contacts.

### Model calibration

We calibrate the model to fit the basic reproduction number *R*_0_ = 2.78 obtained by fitting the cumulative number of cases with the SEIR model adjusting the model hyperparameters: *r*, *T* and *p_c_* for labeling a contact as infectious. For this calibration we trace contacts among the COVID-19 patients. We use a window of 4 days before and 7 days after the onset of symptoms which is the extended infectious period from Figure 1a. The detected contacts are labeled as infectious if the target starts with symptoms within a maximum of 14 days from the exposure. Only the first contact that fulfills the requirements is considered infectious. After running the calibration process we find the hyperparameters of the contact model that fit the *R*_0_ estimated value.

The calibration period runs over the month of March, 2020 in the state of Ceará. In March 2020, there were 1392 infected cases reported in the state dataset localized in the city of Fortaleza. Out of these cases, we cross-checked 465 infected users in the GPS dataset. We then trace the contacts of each 465 infected users over the infectiousness period of -4d/+7d from date of first symptoms against the remaining 464 infected users. We run the calibration over a set of hyperparameters to search for the best set that most closely approximates the basic reproduction number *R*_0_ = 2.78. The closest fit is obtained for: *T* =30 min, *r* =8 m, and *p_c_* = 0.9.

Using these hyperparameters, we find that 90 unique infected users participated in contact events with other infected users. That is, 90 users have either a non-zero in-degree or out-degree or both. The remaining 375 users had neither in- nor out-degree detected in the GPS database. The 90 users are plotted in the transmission tree in Fig. 1c. The tree contains 60 contact events. That is, the total number of out-links (and in-links) in the tree is 60. Out of this 60 contacts, some of them coincide to the same target (see Fig. 1c). Since a target can only be infected once, we do not count these duplicates to find 52 unique contacts. This results in an average out-degree for the set of located infected users of 〈k_out_〉 = 52/465 = 0.112. Since the calibration is run with a GPS dataset sample of 111,168 users out of the total population in the city of Fortaleza of 2,643,247, then we scale the average out-degree by a factor of 23.77 and obtain a calibrated 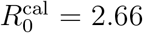 which is in accordance with the estimated 2.78 value. Thus, the hyperparameters are set to *T* = 30 min, *r* = 8 m and *p_c_* = 0.9.

The distribution of probabilities *P_i_*[*n*] from Eq. (1) shown in SI Fig. 11a appears to be extremely polarized leaving the *p_c_* with a wide range of optimal values for an appropriate fitting. We use *p_c_* = 0.9 which fits our results and at the same time is large enough to filter irrelevant noisy contacts (the results do not depend largely on *p_c_*). As shown in SI Fig. 11b, *T* = 30 min is also the value at which the average of the contact probability 〈*P_i_*[*n*]〉*_T_* starts to decrease with *T*. Therefore, *T* = 30 min is the smallest value of a well-behaved *T*, since we expect that the contact probability should decrease with *T*. Eight meters is also consistent with typical precision of geolocation given by noise level.

### Percolation, giant connected component and centralities

In network theory [18], the percolation problem studies the dismantling of networks under removal of nodes or links (bond percolation), as well as associated problems of disease transmission [9, 10], robustness and resilience of networks [11, 12]. The GCC is the largest connected subgraph of a graph, i.e., in the GCC there is always a pathway to reach a node from any other node. Formally, the network dismantling occurs at a critical percolation transition threshold *q_c_* of the fraction of removed nodes *q*. The GCC occupies a fraction larger than zero for *q* < *q_c_* and vanishes otherwise. The vanishing of GCC at *q_c_* marks the percolation transition between two phases, namely, a connected phase and a disconnected one.

One can remove nodes or links at random [9, 18] or by following optimized strategies to break the GCC with the minimal number of removals. We study different strategies to destroy the GCC with minimal removals based on removing nodes in the network by ranking them according to different centralities. All strategies are adaptive, meaning that the ranking is recalculated after every removal. We use:

1. Degree strategy: We rank the nodes by their degree (number of contacts) from top (hubs) to low degree [11, 12] and then remove nodes starting from the hubs adaptively.
2. K-core strategy: We rank the nodes by their occupancy in the k-shells of the network. The highest rank corresponds to the inner k-shell, that is the maximal k-core at 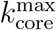. Since each k-shell is formed by many nodes, we then rank the nodes inside a given k-shell by their degree. We then remove each k-shell in turn, always recalculating the ranking after every removal [22].
3. Collective Influence (CI) strategy: we calculate the CI of each node according to Ref. [13] with ℓ = 3 and remove the nodes with CI adaptive.
4. Betweenness Centrality (BC) strategy: We rank the nodes by their BC and remove them one by one from highest to lowest, adaptively [26, 27, 29]. The betweenness centrality of a node is proportional to the number of shortest paths that pass through the node [26, 27, 29]. It is calculated by considering all the pair of nodes in the network and calculating the shortest path between each pair. This methods was found in previous simulations to be a good predictor of a node’s epidemic influence in a contact network [28]. For larger datasets used in this study, approximate fast algorithms can be used to calculated BC. See Ref. [32].
5. We also use other strategies, like eigenvector-based centralities and combinations of other centralities to characterize the node importance [33].

We test the most efficient strategy to dismantle the transmission network. We plot the normalized size of the GCC, *G*(*q*), that is, the number of nodes in the GCC after removal of a fraction of q nodes divided by the size of the GCC at *q* = 0. As nodes are removed from the network, we search for the strategy that provides the minimal removal with the maximal damage to the GCC. The best strategy over all the networks studied across all of the above ranking is the high BC strategy.

### K-cores and k-shell decomposition of the network

The k-core of a graph is the maximal subgraph made of nodes with degree *k* or more. A k-shell of a graph is composed by all the nodes that belong to the k-core but not to the (*k*+1)-core. See SI Figs. 12 and 13 for definitions in a network with 3-shells, i.e., with a maximal 3-core 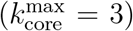, and Fig. 14 for an example in a real network.

The k-shell decomposition assigns each node to a k-shell in the network. The k-cores are nested structures and k-shells are disjointed; e.g., the 2-core contains the 3-core and so on, and the 2-core is formed by the 3-core plus the 2-shell. By definition, the GCC is the largest 1-core. The maximal k-core is the inner subgraph of the network and it is indexed by 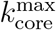 index. The low k-shell are the peripheric shells.

In practice, the k-core of a network is obtained by iteratively removing all nodes with degree smaller than *k*. One starts the removal process by removing all nodes of degree one (see Fig. 13). After the first removal, nodes that initially had degree larger than one may end up with degree equal to one. Then, one repeats the process until no remaining node in the network has degree equal to one, or equivalently, every node in the network has degree at least equal to 2. This set of nodes with *k* = 2, or higher, is the 2-core. The other k-cores are obtained in analogous manner.

It is important to note that a k-core can be composed of disconnected clusters or components. For instance, the example 3-core in Fig. 13c contains 3 components. These three components are disconnected in the 3-core, but they are integrated in the network by nodes belonging to the 2-shell as shown in Fig. 13a. This is an important property of the k-cores found in the transmission networks during the lockdowns. This property is seen in the networks studied in the main text. For instance, the network of Ceará from March 27 plotted in Fig. 3b has a rich k-core structure shown in Fig. 14 where, for instance, the 6-core is composed of 5 disconnected components. This is an important property for an strategy based on betweenness centrality. Typically, we find that these components are joined together by nodes in lower k-shells, which are identified by their high betweenness centrality. Then, the k-cores components can be relatively easily dismantled by a few removals outside the k-cores.

Notice that the maximal k-cores are composed of nodes with high degree that connect with other nodes of high degree, which in turn also connect with others of high degree, and so forth. This implies that the k-cores do not have dangling ends made of nodes with degree smaller than *k*. That is, the k-cores are close, in a sense. The k-shell decomposition then cleans the network of those low degree dangling ends in a systematic way and reveals the core of the network, which is the most important part for spreading. See an interpretation of k-core in terms of ecosystem stability at Nature Phys. 15, 95-102 (2019).

Notice also that a hub can be in the maximal k-core or in an outer k-shell according to how the hub is connected. For instance, the red hub in Fig. 13a is in the 3-core because it is also connected to other hubs with 3 or more connections. However, the orange hub is in the 1-shell in Fig. 13a because it is connected with nodes with low degree.

Monitoring the k-cores of the networks as a function of time we find that before the lockdown the maximal k-core index is in average around 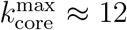 and then drops to half of this value with a maximal 6-core in average during the lockdown 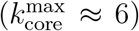. SI Fig. 16 shows this drop in the maximum k-core index from 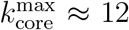 to 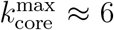, in average. Since 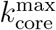 changes with time, to study the occupancies of the k-cores and k-shells across the quarantine transition, we define the *∊*–kcore as the k-core with *k* such that 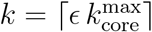. The complement of the *∊*-kcore is the *∊*-kshell defined as the union of the remaining k-shells with k such that *k* = 1, 2,…, 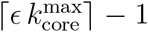. Thus, the union of the *∊*-kcore and the *∊*-kshell constitute all the network. In the paper we consider *∊* = 0.5 that divides the k-shells in two.

## SUPPLEMENTARY INFORMATION

We include Supplementary Information Figures and Section I on Sampling bias, robustness and incompleteness of the dataset.

### I. SAMPLING BIAS, ROBUSTNESS AND INCOMPLETENESS OF THE DATASET

This section addresses concerns regarding the extent to which the present results would hold under implementation in terms of robustness to data quality and coverage, sampling bias on demographics such as coverage of location, socio-economic status, age and gender and privacy. Some of these issues are discussed elsewhere [28, 34].

#### A. Sampling bias

Uniformity of data coverage of the dataset across socio-economical classes, age groups, and geographical regions may affect our results due to the relatively low coverage of the GPS data. This problem is important given that the sample of the population is relatively small compared to the underlying population: 111,168 users out of the total population in the city of Fortaleza of 2,643,247 at 4% of the population and the biases could be substantial. Further, this problem is important for COVID-19 which itself has non-uniform incidence across these factors. Thus, we have investigated the sampling bias of our GPS population https://en.wikipedia.org/wiki/Sampling_bias.

We investigate the most likely bias dominated by geographical coverage, socio-economic status (wealthier over-represented) and age and gender of users (younger over-represented) since one would expect the majority of symptomatic cases are in the lower end of the socio-economic spectrum, and older age groups. We quantify these biases in the GPS dataset from apps by assigning each mobile app user to a geolocalized residential area defined as the place where the user spends most of the time at night between the hours of 10 PM and 5 AM in the period of study. Using these data we study the distribution of geographical localization of the app users. We consider the 120 neighbourhoods (quarter or ’bairro’ in Portuguese) defined by the administrative boundaries in Fortaleza. Since the neighborhoods are extensive, the geolocalization of the users is non-identifiable. The population of each neighbourhood is provided by the Instituto Brasileiro de Geografia e Estatistica (IBGE or Census Bureau). By using the geolocation of each GPS user we calculate the fraction of app users in each neighborhood and then compare with the real fraction of the population of each neighborhood obtained from IBGE. The distribution of the populations obtained from GPS data and the real population distribution from IBGE are shown in Fig. 5. We perform a two-sample Kolmogorov-Smirnov (KS) test and find p-value =0.388, KS distance = 0.117, indicating that we cannot reject the hypothesis that the GPS data and the real data come from the same distribution. Therefore, we conclude that the GPS data has an acceptable geographical coverage of the real population indicating no sampling bias in geolocation under a statistical test.

**FIG. 5.**
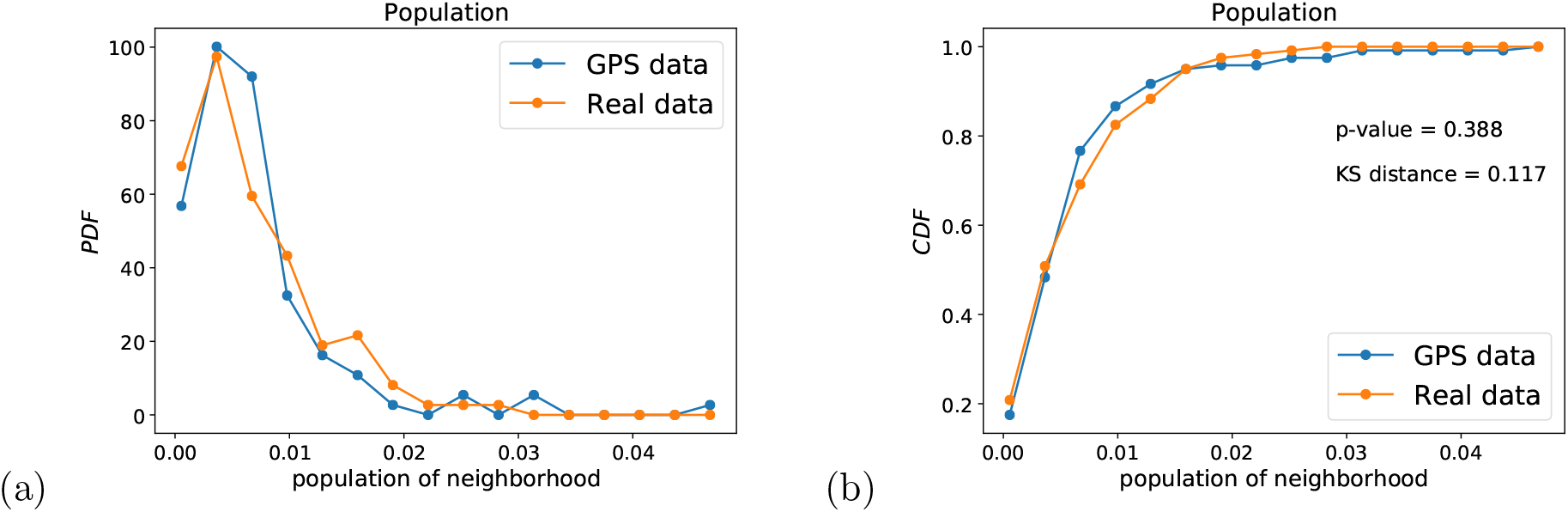
(a) Probability density function and (b) Cumulative distribution function of the fraction of the population per neighborhood in Fortaleza to the total population. We show the real distributions and the distributions from the apps GPS data. Both distributions pass a two-sample KS test indicating that we cannot reject the hypothesis that they come from the same distribution under the test.

Furthermore each neighborhood has a distinct Human Development Index (0 <HDI< 1) provided as well by IBGE. By using this metric of socio-economic status, we study the possibility of socio-economic bias in the population sample. We now cluster the neighborhoods by their HDI and plot the PDF and CDF of the population of neighborhoods (measured as the fraction to the total population) with a given HDI obtained from the GPS data sample and from the real population from IBGE. Results are shown in Fig 6. We performed a two-sample Kolmogorov-Smirnov test and find that the sample distribution of HDI obtained from the GPS data and the real population distribution of HDI socio-economic status pass the KS test, p-value = 0.699, KS distance = 0.250. Tus, we cannot reject the hypothesis that the GPS and real data come from the same distribution indicating lack of sampling bias in socio-economic status under this statistical test.

**FIG. 6.**
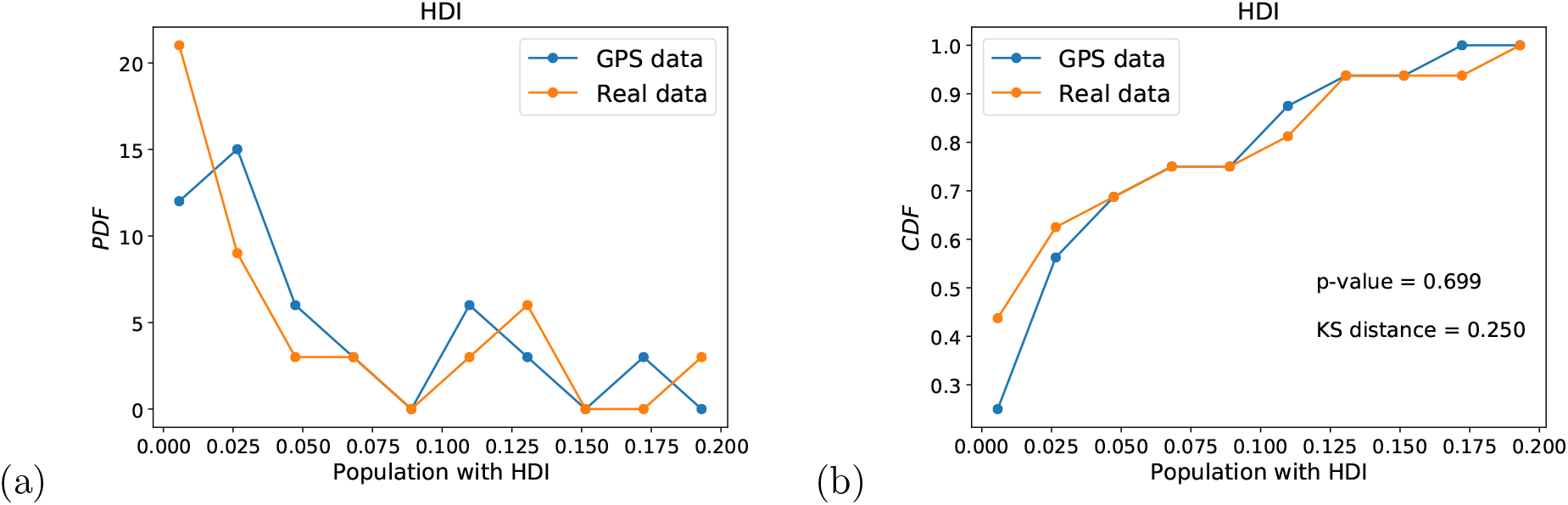
(a) Probability density function and (b) Cumulative distribution function of the fraction of the population per neighborhood with a given HDI in Fortaleza to the total population. We show the real distributions and the distributions from the apps GPS data. Two-sample KS test indicates that we cannot reject the hypothesis that the real and GPS sample come from the same distribution under the test, indicating lack of sampling bias under this test.

The two final tests of sampling bias are done on the age distribution and the gender distribution. We do not have direct access to the age and gender of the GPS users. However, an indirect test of sampling bias can be used using the patient data. We compare the distribution of age and gender in the full patient dataset with the distribution of age and gender of those patients that are localized in the GPS dataset. The hypothesis is that if the GPS dataset is biased by age or gender (for instance, if the app users over-represent younger people) then the distribution of localized patients in the GPS dataset should reflect this bias respect to the distribution of age and gender of the whole patient population. Figure 7 shows the respective PDF and CDF. We find that we cannot reject the hypothesis that the CDF of age from GPS data and the patient data come from the same distribution with p-value = 0.785 and KS distance = 0.039, indicating good coverage of age distribution (Fig. 7a, b). The distributions of gender shown in Fig. 8a, b indicate also the lack of bias in the gender distributions.

**FIG. 7.**
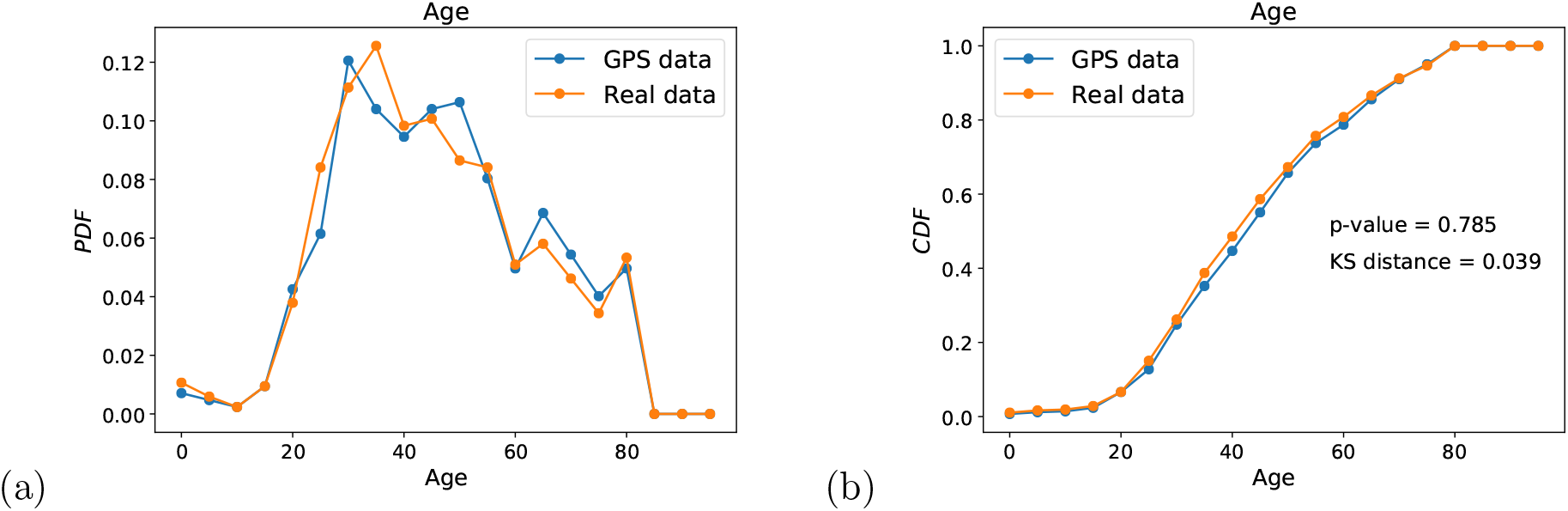
(a) PDF and (b) CDF of age distribution in the GPS geolocalized data compared with the real patient data. We cannot reject the hypothesis that both samples come from the same distribution under KS statistical testing.

**FIG. 8.**
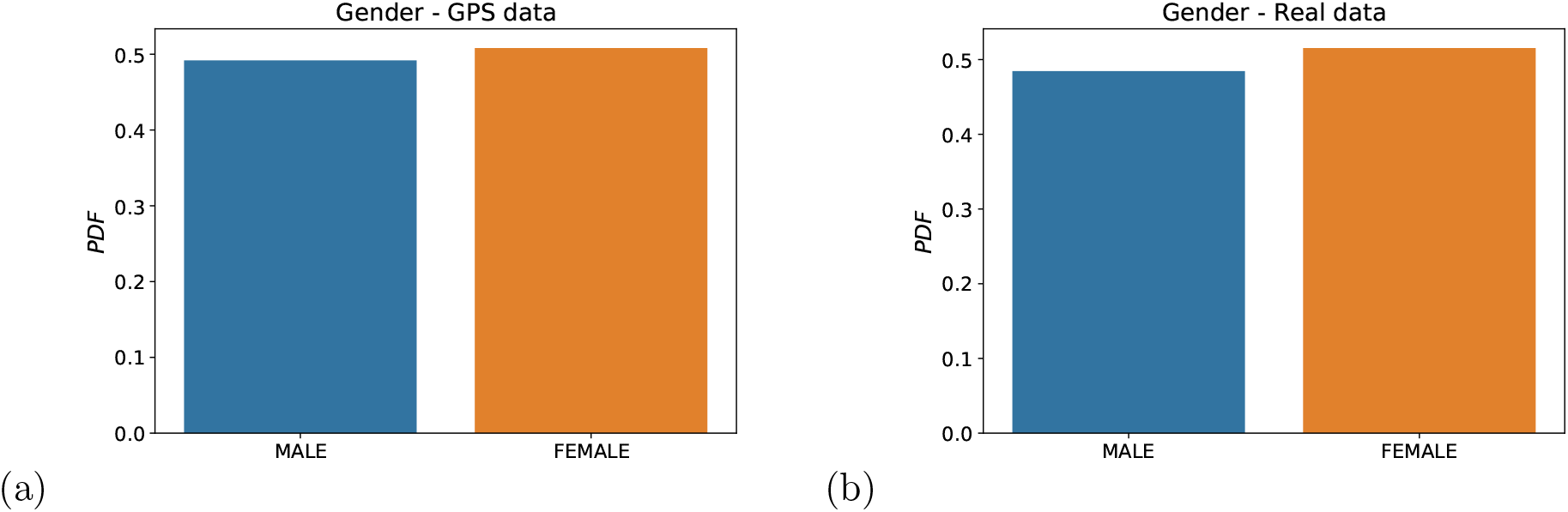
(a) PDF and (b) CDF of gender distribution in the GPS geolocalized data compared with the real patient data suggesting lack of bias.

We conclude that the GPS sample does not have significant bias under statistical testing in the considered demographic variables.

### B. Correlation between a unique mobile ID and a unique person

To test this correlation we investigated whether the geolocalized patients in the GPS datasets have (1) visited any hospital (near 200 m) or (2) have visited any pharmacy on the date of test results. We find that out of the 1,440 patients identified in the GPS dataset 1,224 (85%) have been identified with the second test.

### C. Uncertainty in the first day of symptoms

The day of first symptoms is the date reported retrospectively by the patient of appearance of symptoms at the first consultation at a healthcare facility, thus, there is some uncertainty in its determination related to the patient’s report of this date. This uncertainty could affect the results of the contact tracing. For proper functioning of the algorithms, data should be fed into the algorithm of contact tracing in real time, while for most of the cases the first date of symptoms is reported retroactively. In a mobile app implementation of contact tracing, the user should be given the capability to report the symptoms in real time as soon as they develop, via the app, thus diminishing uncertainties in the proper definition of the window of observation to detect contacts.

### D. Temporal sampling

The distribution of temporal sampling of GPS ping datapoints per user in our GPS dataset displays four peaks: around zero, at 5 minutes, 10 minutes and 20 minutes, see Fig. 9. This is consistent with other apps using, e.g., the Google-Apple framework. Typically, this is a trade-off between accuracy and battery life. In our data, pinning distributions are uniform across day and night. We have investigated the robustness of our contact tracer to different ping intervals. The effect is particularly important since betweenness centrality and k-core are both macroscopic properties, meaning a small change in the network can create large changes everywhere in the network.

**FIG. 9.**
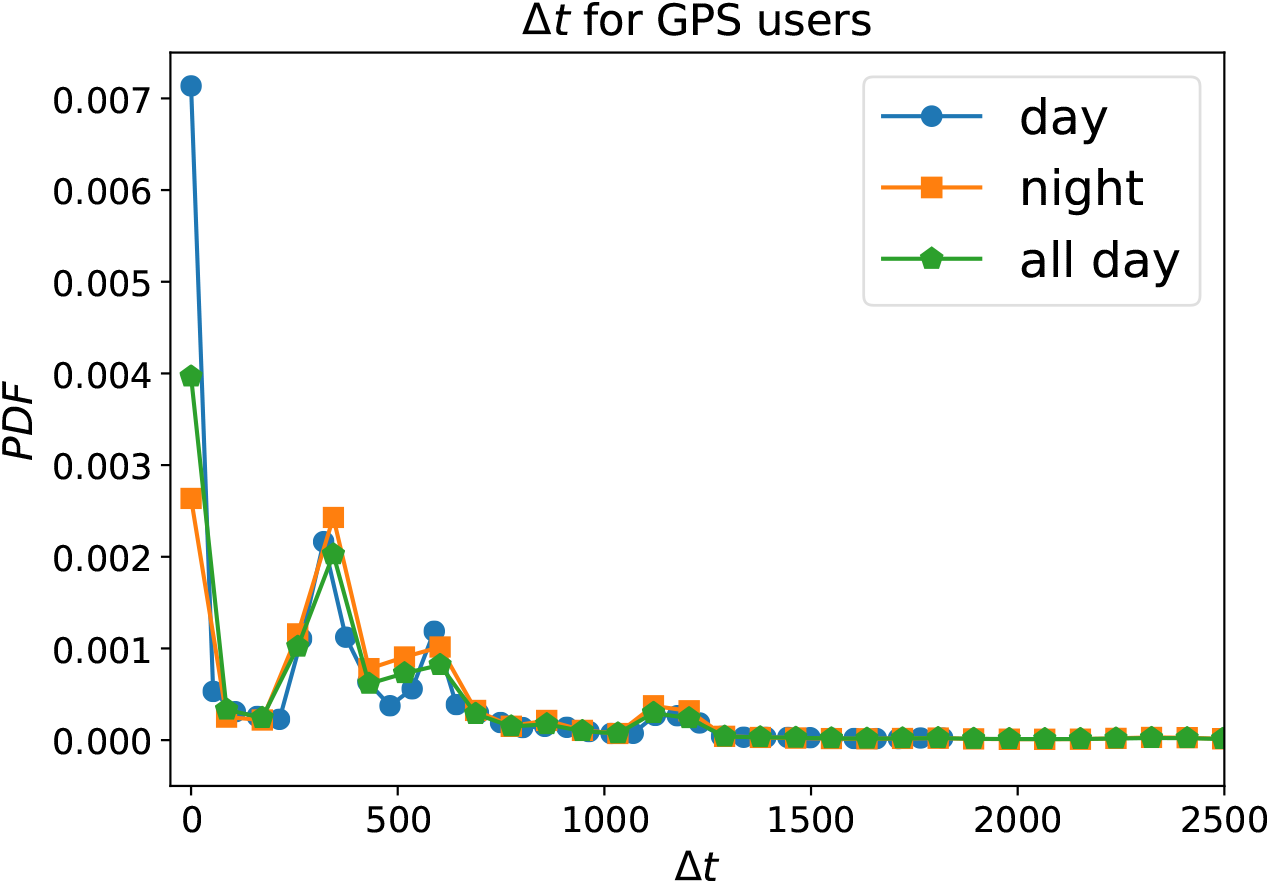
Distribution of the time interval between GPS pings during all day and separated by day and night.

**FIG. 10.**
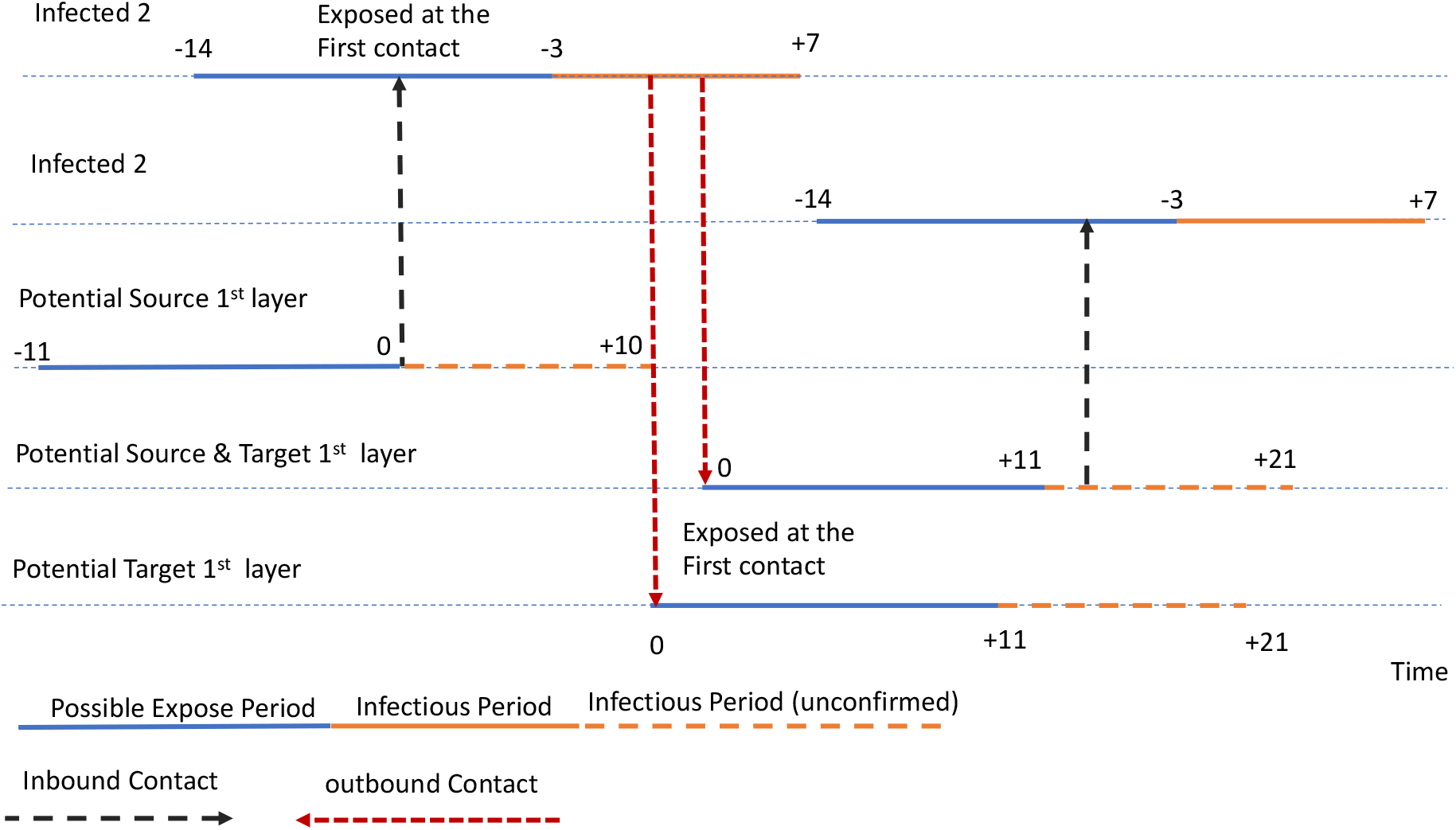
Contact, layers or pre-symptomatic and asymptomatic captured by the model. Our treatment of asymptomatic cases is to increase the exposure period to -14 days to accounting for possible two-chains of infection as shown in the figure. Contacts between -2 days to -14 days from the day of first symptoms are more likely to be an exposure from an asymptomatic infected person. Contact from -2 days to +7 days from first symptoms are considered to be transmissions contacts from the patient.

Figure 11b shows that the a minimal time interval that captures the correct behaviour in the probability to find a contact is around 30 minutes. This is seen in Figure 11b where the correct decreasing behaviour of < *P_i_*[*n*] >*_T_* with *T* appears after *T* > 30 minutes. Notice that this probability is supposed to decrease with increasing *T*. Considering *T*=30 minutes as a minimal time interval to find a contact, we would need at least two pings inside 30 minutes to properly define an interval of contacts. Therefore, we do not recommend to use longer ping intervals than 15 minutes, in order to have enough statistics to capture contact points.

**FIG. 11.**
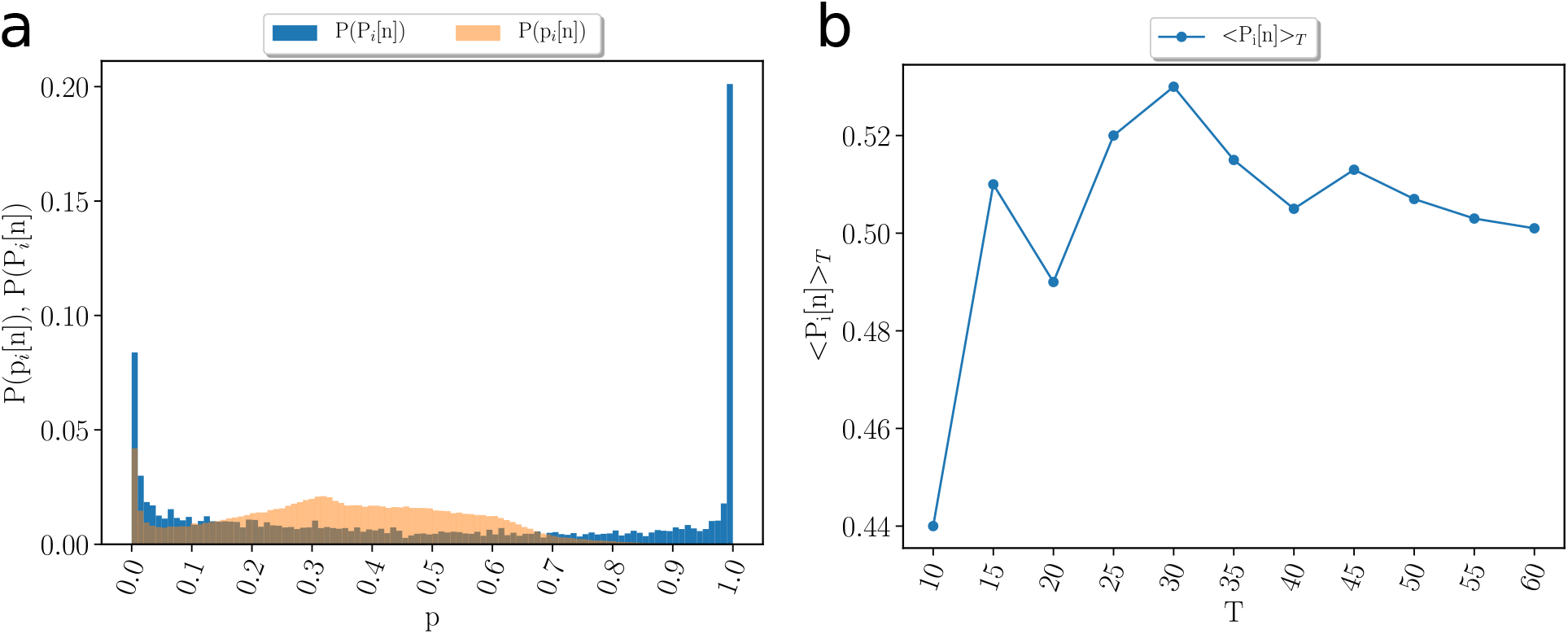
(a) Probability distribution of *p_i_*[*n*] = *p_d_*[*n*] · *p_t_*[*n*] (orange) and the recursive form *P_i_*[*n*] defined in Eq. (1) (blue). The *P_i_*[*n*] are polarized to 0 and 1 becoming the best thresholded metric to use to consider a contact as infectious. (b) Average value < *P_i_*[*n*] >*_T_* as a function of the time window *T* of the spatio-temporal contact area. *P_i_*[*n*] has a peak at *T* = 30 min; it decreases for *T* > 30 min and increase for *T* < 30 min as a function of *T*. The decreasing behaviour is what is expected, thus, 30 min is the minimum bound for the correct value of *T*.

**FIG. 12.**
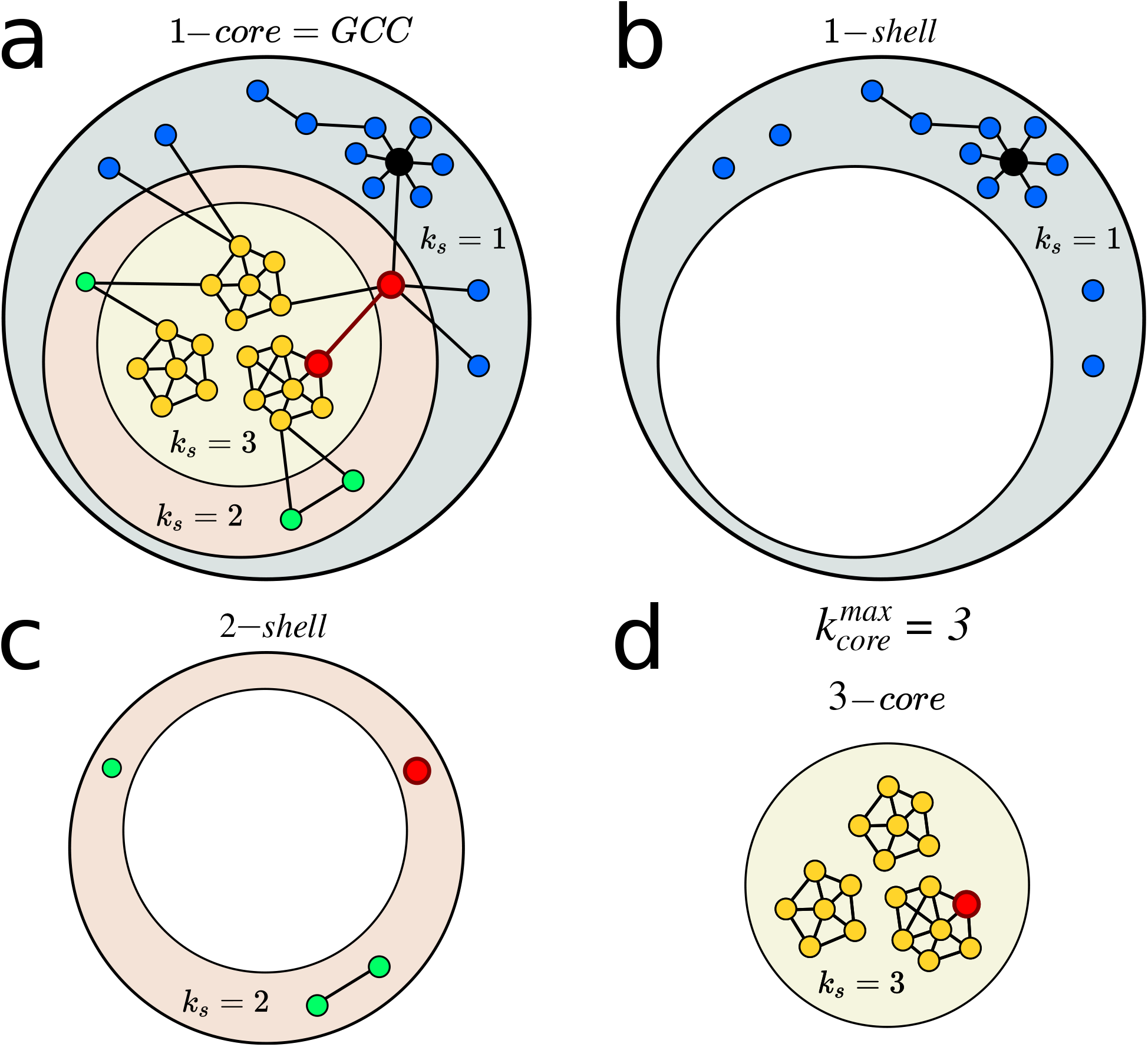
Network structure under k-shell decomposition. **a**, A sample network with 3 shells. The k-shell index *k_s_* is not necessarily associated with other centralities. Here, the hub of the network in black with *k* = 7 is in the 1-shell, *k_s_* = 1. The two top node in betweenness centrality, highlighted in red, belong to the 2-shell and the 3 shell, respectively. The 1-core is equivalent to the GCC. **b**, The nodes with *k_s_* = 1 form the 1-shell, **c**, the nodes with *k_s_* = 2 form the 2-shell, and **d**, the nodes with *k_s_* = 3 form the 3-shell which is also the 3-core.

**FIG. 13.**
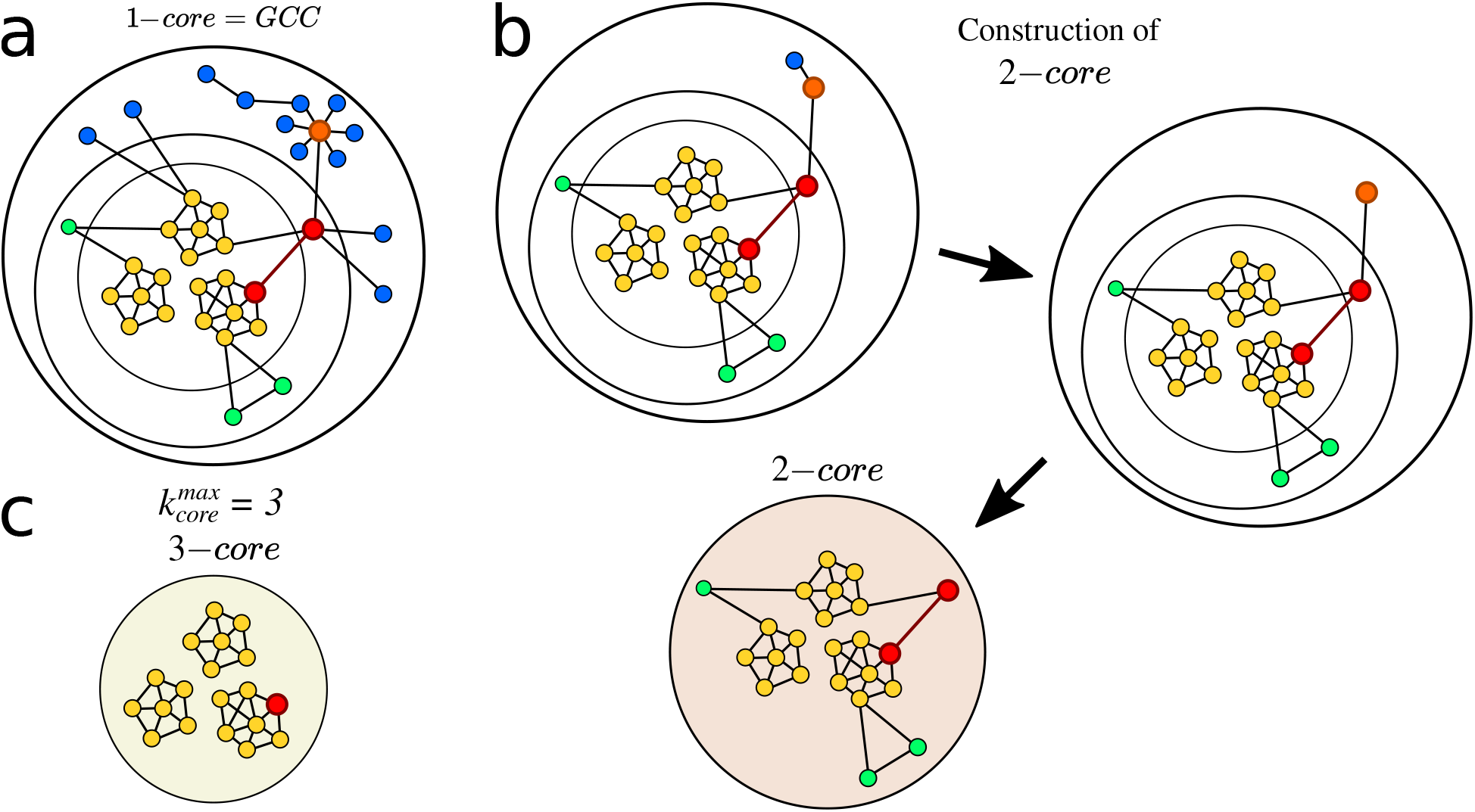
K-cores of a network. **a**, We start the k-shell decomposition with a network configuration where every node has at least degree *k* = 1. This set of nodes forms a 1-core. **b**, Then, every node with *k* = 1 is iteratively removed to obtain the 2-core. As one can see, the removal of these nodes changes the degree distribution. Thus, nodes are removed until all remaining nodes are left with *k* ≥ 2. **c**, Following the k-shell decomposition nodes are removed until we obtain the 3-core. The 3-core can be made of multiple disconnected clusters.

**FIG. 14.**
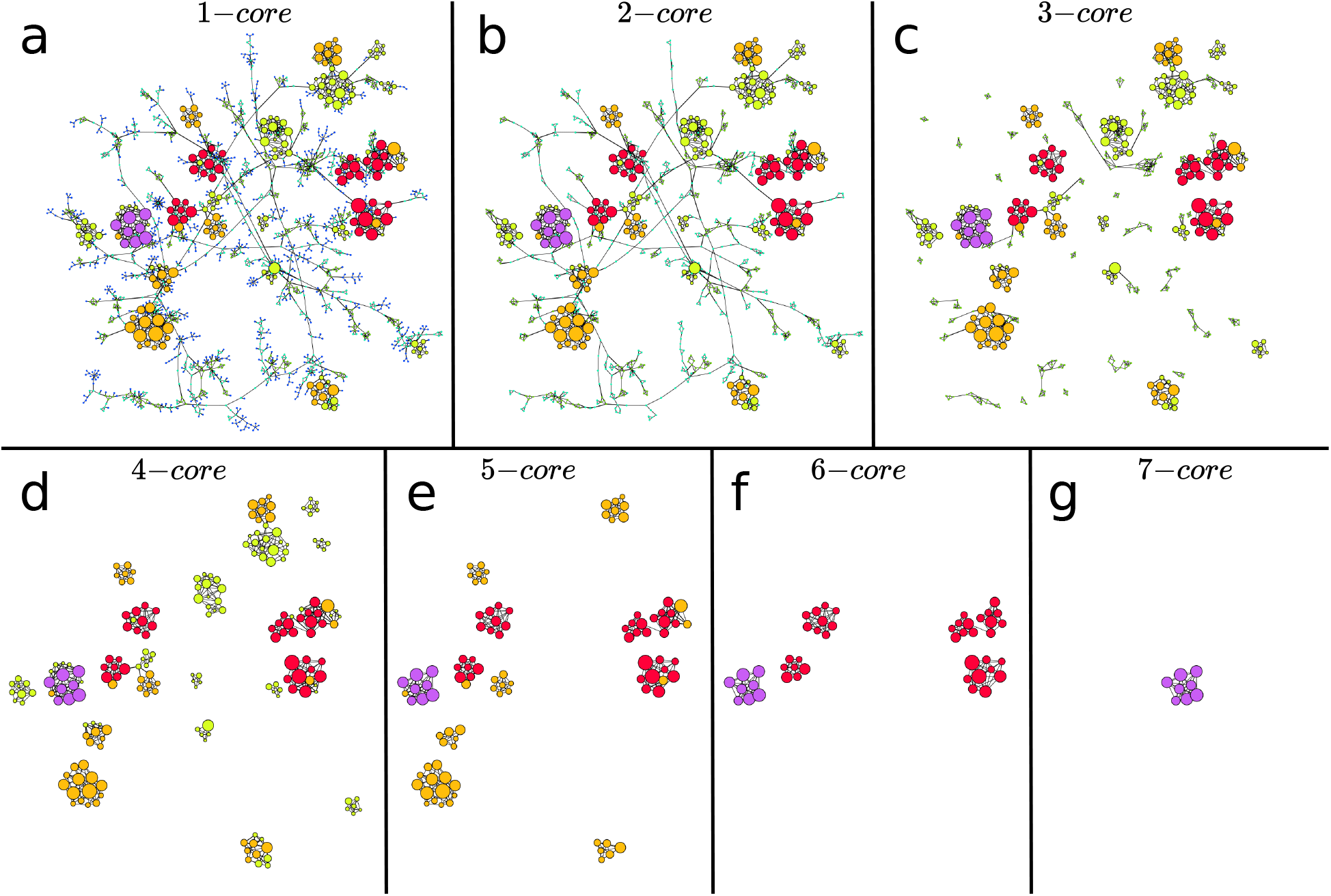
Example of k-core and k-shell structure in the network plotted in Fig. 3b obtained during the lockdown. Here the colors are set by the k-shell occupancy of each node.. Each k-core is composed by the k-shell plus the (k+1)-core. The k-cores are nested structures. For instance, the 5-core in (e) is composed by the 5-shell (yellow nodes) and the 6-core, which, in turn, is composed by the 6-shell (in red) and the 7-core (in purple). Since the 7-core is the maximal k-core, 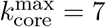 for this network, then the 7-core is also the 7-shell. In this network the 0.5-kcore is the 4-kcore and the 0.5-kshell is composed by the 1-shell plus the 2-shell and the 3-shell. We notice how a given k-core can be composed of many disconnected components. For instance, the 6-core is composed by 5 disconnected components. This is important, since each component of a given k-core can be localized in different areas, like different hospitals, in the map, see for instance, Fig. 3c and 3d. It is also visually apparent that to destroy this network, a direct ’attack’ to the high k-cores is not optimal. Instead, removing the high BC nodes that populate the lower k-shells is the best strategy. We plot each k-core in turn: **a**, 1-core, **b**, 2-core, **c**, 3-core, **d**, 4-core, **e**, 5-core, **f**, 6-core and **g**, 7-core.

**FIG. 15.**
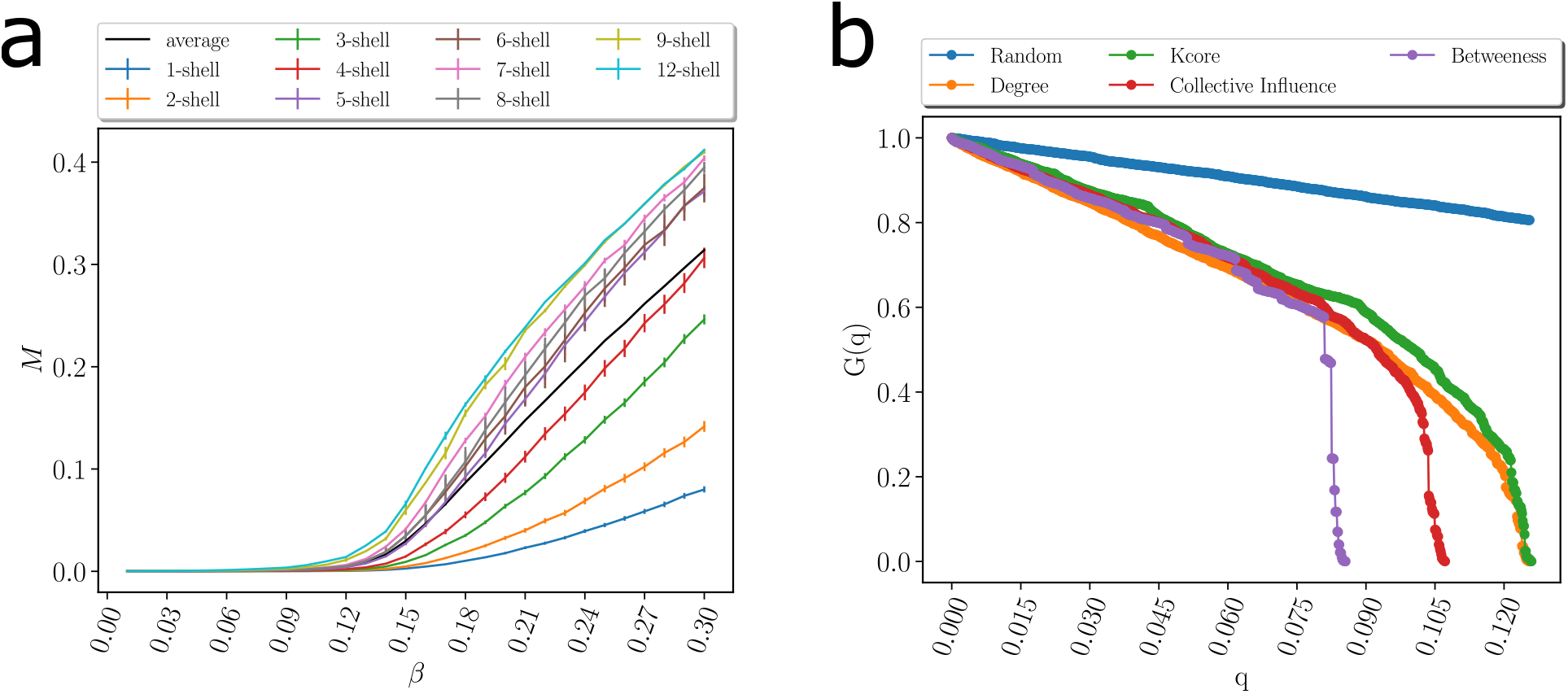
**(a)** Amount of infected population 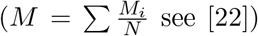 when the spreading starts in a given node in a k-shell as a function of the probability of infection *β* for a SIR model on the same network on March 19 in Fig. 3a. in pre-quarantine Ceará. The black is the average value over all the starting nodes in the network. The average divides the shell contribution to the spreading of the virus in two groups above and below the average. The 0.5-kcore composed of the 6-core (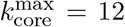 in this network) which contains nodes from the 6-shell to the 12-shell, has maximal spreading. the 0.5-kshell which is composed by the remaining shell from 1-shell to 5-shell has minimal spreading, below the average. **(b)** Optimal percolation analysis performed over the network in Fig. 3a before the quarantine on March 19 in Ceará with different attack strategies and their effect on the size of the largest connected component G(q) versus the removal node fraction, *q*. Depending on the strategy nodes are removed: randomly (blue), by the highest value of betweenness centrality (green) [26, 27], degree (orange), collective influence (red) [13], and by the highest k-shell followed by high degree inside the k-shell [22]. After each removal we re-compute all the metrics. The optimal strategy is removing the nodes directly by the highest value of betweenness centrality.

**FIG. 16.**
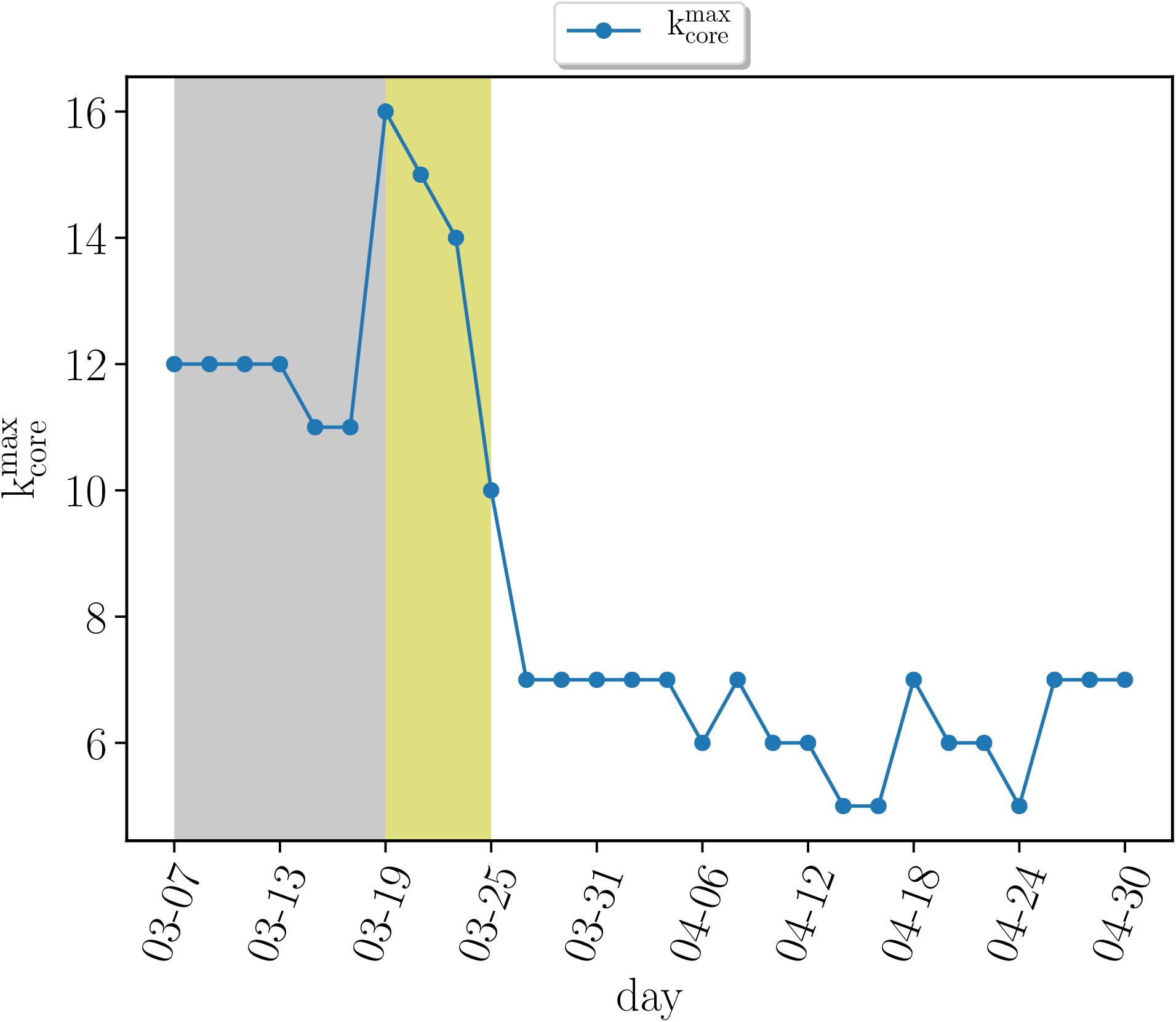
Evolution of maximum k-core index 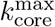 versus time previous to the quarantine (grey area), right after the quarantine (yellow area) and later. We see how the maximum k-core index drops drastically after the mass quarantine.

**FIG. 17.**
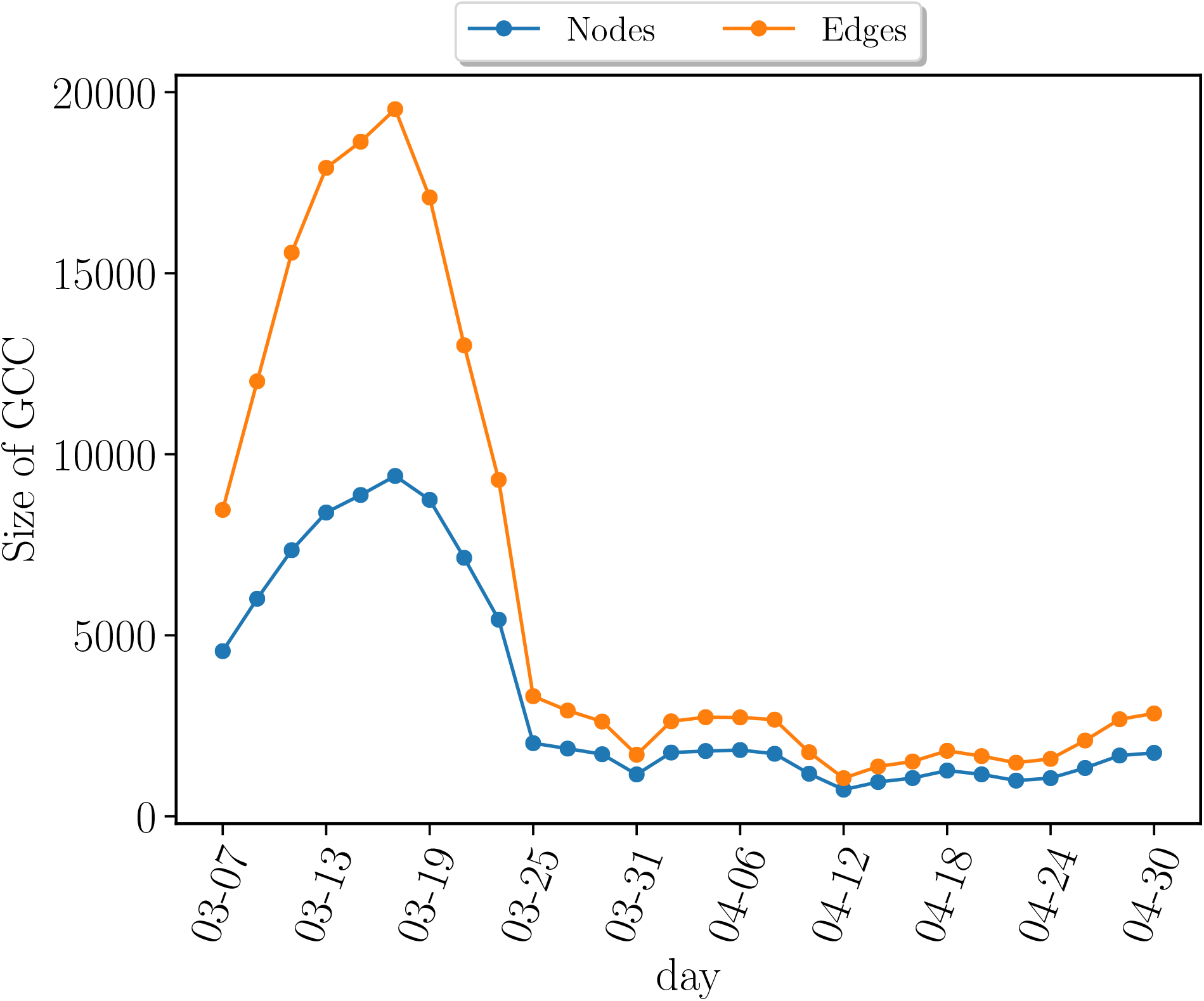
The number of nodes (blue) and edges (oranges) in the GCC versus time. The initial increase in the number of nodes is artificial due to the fact that we perform contact tracing 14 days back for each patient and our data collection started in March 1. Thus the networks in the first two weeks have relatively lower contacts than the rest.

**FIG. 18.**
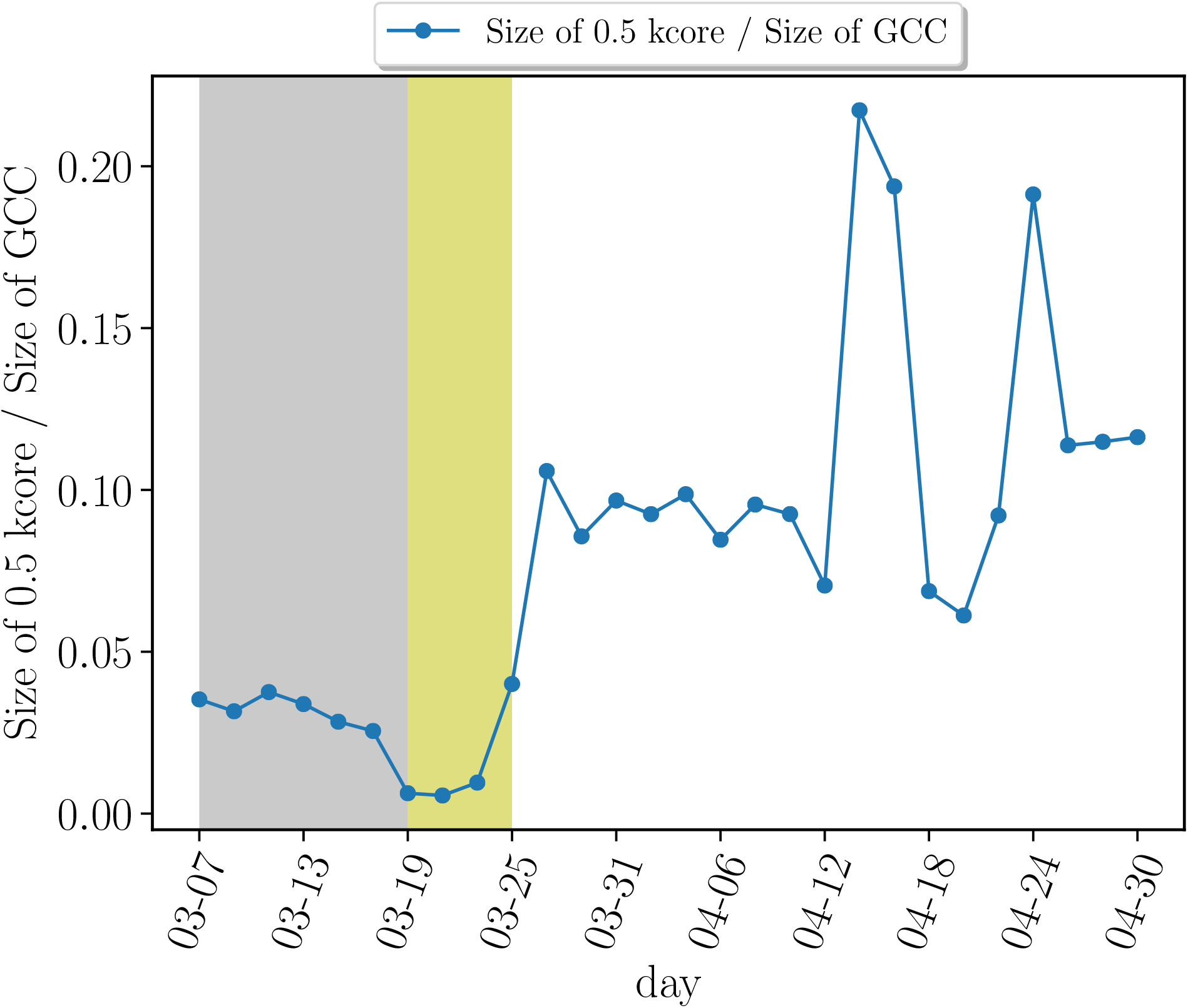
Evolution of maximum 0.5-kcore size versus time normalized by the size of the GCC. The proportion of these maximum k-cores keeps increasing after the quarantine.

### E. Temporal features

Bias in the temporal features of the model. The periods of infectionness and exposure are relaxed conservatively as explained in the text. We studied the robustness under the definition of these periods. We have relaxed the window forward in order to be symmetric with the relaxation of time before and after the symptoms by using 8 days after symptoms and find that the results do not change. This is expected since patients are expected to be either at home in quarantine or in hospital, in average, 5.5 days (95% CI 4.6 - 6.6 d) after first symptoms, according to [17]. Indeed, we find that the displacements of patients are highly reduced after the days of first symptoms. Thus, we expect that the majority of contacts are established before symptoms, highlighting the necessity of contact tracing back in time.

### F. Unmatched cases

There are uncertainties in the number of cases matched between datasets due to the fact that only a fraction of patient cases can be matched to the GPS dataset. The unmatched cases cannot be matched at random to complete the data since these would ignore the correlations between the disease and behaviour. Thus, we do not consider the unmatched cases in the contact network. This unmatching is due to the incomplete coverage of the GPS dataset respect to the real population. However, we have checked in Section IA that the sampling coverage of the dataset is consistent with the real population which then minimizes the chances of small sampling bias. The spreading rate in our dataset is 0.112 as described in the Methods Section Model Calibration. This value corresponds to 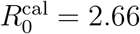 consistent with the real *R*_0_ value in the whole population.

### G. Sparseness of the dataset

Due to the sparseness of the GPS data, we do not have access to all the contact between the infected people. To account for the smaller coverage of the GPS data in the calculation of *R*_0_, we first obtain an effective 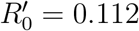 valid for the smaller GPS dataset. This number obtained for the GPS dataset should be rescaled by the population ratio between the real population and the GPS sample, which is a factor of 23.77 and provides *R*_0_ = 2.66, which is consistent with the values directly measured from the data.

It is important to note that this rescaling is only used to estimate the epidemiological parameter *R*_0_ and does not in principle affect the further modeling of the contact networks, except for the fact that we use this estimated value 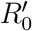 to estimate the hyperparameters of the model (*T*, *r*). This means that, using other GPS datasets with different coverage, a new set of parameters needs to be determined to fine tune the model to the particular coverage of the GPS dataset used. Therefore, in a less sparser dataset in an actual application with larger number of mobile app users, the parameters of the model (*T*, *r,p_c_*) should be calibrated accordingly.

We note that in our rescaling, we do not rescale the network, nor the in- or out-degree distributions, but just use the smaller GPS sample to obtain the hyperparameters of the model.

### H. Time-evolving weak ties

Weak ties in temporal networks have been investigated in [35]. In our case these weak ties are evolving with time as well. While we apply our definitions of centrality metrics to static networks defined over 15 days, we employ a moving window that calculates a new network every three days, thus extending the static definition of weak ties to temporal data.

### I. Infected cases

We have based our modeling on the evolving number of new cases, which is not as robust estimate as the number of death cases. The main indicator in contact tracing is the date of first symptoms. In the absence of this datapoint, the day of hospitalization can be also used to estimate the date of first symptoms from the hospitalization data using the ensemble average of the time interval from first symptoms to hospitalization across the rest of the patients. While death is a more accurate metric than these two metrics, an estimate of the day of first symptoms is more difficult to obtained from the date of death, which anyways, occurs to only a fraction of the patients.

### J. Contact tracing methods

In a contact tracing app based on GPS technology, a centralized server is needed where one entity has access to the GPS data. The algorithms discussed here are also feasible in contact tracing platforms using Bluetooth technology, as long as the detected neighbours are shared at a given point by a central server. This is because, the optimal tracing strategy requires the contacts of the contacts to build the network. On the other hand, the geolocalization of the red zone of contacts in a geographical map with precise locations shown in Figs. 2 and 3, requires the use of GPS data and cannot be performed with Bluetooth-based technology.

### K. K-core infection

We mention an important point about the definition of the k-cores. Given a network, the same k-core can be composed of several disconnected components. This is what we see from Fig. 4c and d, or for instance, the example 3-core in Fig. 13c contains several discon-nected components. Our analyses (displayed in Fig. 4c, d) show that these disconnected components are connected by weak links, which if removed, may isolate the spreading of the virus inside one of the disconnected components of a k-core, containing the spreading of the pandemic to inside the disconnected component of the k-core. In other words, if a part of the k-core is infected, the disease will be controlled within a small group of that k-core and not extended to the rest of the network, if the affected component of the k-core is isolated from the rest by the removal of the weak link.

### L. Intelligent quarantines

It is plausible that the slow decay of cases from the infection peaks after lockdowns observed in many countries might be due, in part, to the lack of deployment of intelligent quarantines based on optimization principles. Our results could help to provide insight into the persistence of infection in many places.

### M. Model of infection

In our model, a transmission probability is calculated for a contact, which is then taken to be infectious if the probability is above a threshold. To avoid overestimating the importance of strong contacts we then define the recursive probability Eq. (1), *P_i_*[*n*], where one strong contact between two highly connected groups is not more important than many weak contacts. This regularization is highly efficient in converting the simple hit probability *p_i_*[*n*] which is observed to be not a good separator of a contact versus a non-contact since its distribution is not bimodal (orange curve) as observed in Fig. 11a, into a bimodal distribution for *P_i_*[*n*] as observed in the figure (blue curve). Thus, *P_i_*[*n*], which takes into account the importance of many small contacts separates well the contacts, and does not need of a precise threshold to be implemented. In fact, any threshold in the range 0.1 < *p_c_* < 0.9 gives similar results in the contact model since most of the cases are concentrated in the extreme cases *P_i_*[*n*] ≈ 1 and *P_i_*[*n*] ≪ 1 or zero. Thus, while the thresholding of *p_i_*[*n*] has a danger of throwing out many weak links, the use of *P_i_*[*n*] regularize this function in the proper way such that the effect of choosing a threshold is minimized. That is, since the distribution of *P*(*P_i_*[*n*]) (Fig. 11a) is highly bimodal concentrating near zero and one, then we were able to separate well a contact *P_i_*[*n*] ≈ 1 from a non-contact *P_i_*[*n*] = 0, and the need for a threshold to distinguish between these two extremes disapears, since the probability is nearly zero between these two extreme values as shown in the figure.

### N. Dynamics of weak links

In principle, the networks are calculated continuously in time, and therefore the weak links are identified as they form and break k-cores. If by removing an individual, somebody else replaces that role, then a new network should identify this new individual added as a new weak link. However, this process may become somehow impractical due to the continuous removal of those weak links. This problem can be treated by determining the roles or occupations of the weak links and then search for a way to remove this risk by targeting those roles or occupations. This can be achieved by the analysis shown in, for instance, Fig. 3c and 3d, by geolocalizing the weak links and k-core in the map to discover the occupations/roles or places visited by the weak links and develop a targeted approach accordingly. For instance, Fig. 3d shows where these occupations and roles contributing to the transmission chain are occurring. Targeting these occupations and places to remove the risk is an efficient way to break the chain of transmission.

### O. Uncertainties in infected population

We match the infected individual by a rule that uses the geolocation of the individual at the address of the patient. This method may cause some uncertainty in the results as some fraction of the infected individuals could be in principle replaced by non-infected individuals. To study this problem, we test that the identified mobile ID has also visited the hospital and or a pharmacy on the date of test results. Using this second test, we corroborated the matching between patients and mobile users. See Section IB.

### P. Level of uncertainty required for implementation

The methods of the algorithm requiere a level of certainty in the dataset that may be not appropriate for the level of uncertainty inherent in the underlying data. The reality of tackling SARS-CoV-2 with digital traces is that data incompleteness may not permit the type of analysis described here. If the data has a large uncertainty, using global network measures such as betweenness to break up the transmission network may not be practicable. To address whether the conclusions are robust we have undertaken study of bias and uncertainty. We perform a treatment of bias, uncertainty and data incompleteness by studying sampling bias in demographic variables like geographical population coverage, socio-economic status, age and gender.

Extensive analysis are reported in Section I A, **Sampling bias**, showing that the distributions of location, economic status, age and gender are similar under two-sample KS test to the distributions of the real data. That is, we cannot reject the hypothesis that the real data and the GPS data comes from the same distribution. Thus, our results suggest that a collection of apps with GPS geolocalization provides a statistically significant sample to study the behaviour of the real population.

Regarding the use of global quantities, such as the betweeness centrality, it may not seen practical, in principle, due to the necessity to obtain the global network. However, according to the Covid Tracing Tracker from MIT Technology Review at https://bit.ly/2Y1NMet, which tracks the contact tracing apps around the world, 35% of government-backed contact tracing apps are based on the same GPS technology used in our study and are able to provide the network of contacts needed for our algorithm to work. These GSP-based apps capture the necessary information on the global contact network needed to perform the present analysis, and our algorithm can be directly applied to them.

### Q. Global measures and scalability

A drawback for the use of a global measure like betweeness centrality is the poor scalability of the measure for large system sizes. However, there are linear approximations of these algorithms that can be used to approximate the metrics for large systems [32]. For larger datasets approximate fast algorithms can be used to calculated BC, see Ref. [32]. Furthermore, once the network is obtained, the chain of transmission can be destroyed by other measures other than BC, which scales linearly with system size and are quite fast to calculate like the degree or CI, although not as optimal as through the weak links as shown by our results.

### R. Asymptomatic cases

Detecting asymptomatic cases is one of the biggest challenge of the COVID-19 pandemic. We have included the existence of contacts with asymptomatic in our model. Our method allows, in principle, the determination of posible contacts with asymptomatic infectious people. As explained in Fig. 1a and further explained in Fig. 10, we extend the exposure period to -14 days from the day of first symptoms. At the same time, the infectious period starts -2 days from symptoms according to [15]. Therefore the contacts identified between -2 days and -14 days (labeled as E’ in Fig. 1a and also as the inbound contacts in Fig. 10) correspond to inbound exposures from asymptomatic carriers. Thus, our model treats asymptomatic cases by considering this exposure period and accounts for possible two-chains of infection as shown in Fig. 10. Since the exposure period with asymptomatic (-14 to -2 days) is longer than the period of infectionness (×2 to +5 days), we obtain a larger number of asymptomatic exposures than infectious transmission contacts per patient.

Detecting asymptomatics is the key to stop this pandemic. Asymptomatics detected at exposure E’ should be immediately tested, even though they have not presented symptoms. Beyond our modeling, we are not aware of other contact methods that have attempted to detect asymptomatic contacts (beyond large-scale widespread testing). Our contact tracing algorithms might be able to detect those asymptomatic transmissions which are critical to stop the pandemic.

### S. Tenuous contacts

Linking infected individuals to GPS traces via space is tenuous and multiple hits are more than likely. To address this problem we first investigated the correlation between a unique mobile ID and a unique patient as discussed in Section I B. Furthermore, to address the multiple hits that are likely to lead to contagion, we have used the recursive probability *P_i_*[*n*] defined in Eq. (1) and explained thereafter.

### T. Quantification of uncertainty in data

Quantification of uncertainty in data plotted in the figures is done via the calculation of the standard error (SD). For the SIR models, the errors are computed as the SE for a given value of the k-core. We notice that some plots are single instances, like for instance the giant connected component, and do not have SE.

### U. Privacy considerations

#### Ethics and privacy consideration of data sharing and using large datasets in biomedical research

The patient datasets were collected by the Epidemiological Surveillance Department, Fortaleza Health Secretariat (CEVEPI) at the Prefeitura de Fortaleza, Ceará, Brazil by the team of Dr. Antonio S. Lima Neto. The present project follows the recommendations of the Wellcome Trust 2016 *“Statement on data sharing in public health emergencies”*, that states: *“In the context of a public health emergency of international concern, there is an imperative on all parties to make any information available that might have value in combating the crisis”* and the statement of the WHO: *“data are the basis for all sound public health actions”*. The Epidemiological Surveillance Department have provided the anonymized data on COVID-19 patients in the City of Fortaleza including the SARS-COV-2 test detection date and first day of symptoms of COVID-19 geocoded cases. Prior to be analyzed, the dataset was completely de-identified at CEVEPI. The data did not include any element that allow us to make a full profile of the patient, such as, the name of the patient, neither the residential address. Additionally, there were no codes associated with the variables that may allow individuals to be identified.

The policy statement for the use of the data states that the information was collected and anonymized by CEVEPI and analyzed by the team at CCNY and UFC, and then deleted completely from the servers of the team at both CCNY and UFC. At the time of submission, the provided datasets have already been deleted from the servers at CCNY and UFC. The data were made available to the team over a period from March 1, 2020 to June 7, 2020 for the purpose of helping the government contain the pandemic. The protocol of contact tracing developed in this study was done within the framework of the public functions of the government aimed at protecting and guaranteeing the public health of the citizens.

The team has signed confidentially agreements stating that the member of the team are responsible for the custody of the data received and that we guarantee the privacy, the confidentiality, anonymization and used of the obtained information of the patients and any third party. The confidentially agreement also states that the team will receive the data only for the purpose of the current investigation and that all data will be erased from the servers at CCNY and UFC at the end of the project in July 2020. All data has been already deleted from the servers at CCNY and UFC and remains under the control of Department of Epidemiological Surveillance, Fortaleza Health Secretariat.

Given the extraordinary nature of this health emergency, it is critical for governments and agencies to share these data with scientists to execute critical studies under the understanding that by using the contact tracing algorithms developed in this study, we could help avoid more casualties during the pandemic. The patient dataset from the Health Department authorities in Fortaleza has the required ethics approval from the IRB at CCNY and UFC. Consent to use these data was given by the Mayor of Fortaleza and the Prefeitura of the City of Fortaleza. The original dataset is kept under the care of the Health Secretariat of Fortaleza.

